# A Novel Model for Simulating COVID-19 Dynamics Through Layered Infection States that Integrate Concepts from Epidemiology, Biophysics and Medicine: SEI_3_R_2_S-Nrec

**DOI:** 10.1101/2020.12.01.20242263

**Authors:** Jack M. Winters

## Abstract

**Introduction:** Effectively modeling SARS-CoV-2/COVID-19 dynamics requires careful integration of population health (public health motivation) and recovery dynamics (medical interventions motivation). This manuscript proposes a minimal pandemic model, which conceptually separates “complex adaptive systems” (CAS) associated with social behavior and infrastructure (e.g., tractable input events modulating exposure) from idealized bio-CAS (e.g., the immune system). The proposed model structure extends the classic simple SEIR (susceptible, exposed, infected, resistant/recovered) uni-causal compartmental model, widely used in epidemiology, into an 8^th^-order functional network SEI_3_R_2_S-Nrec model structure, with infection partitioned into three severity states (e.g., starts in I1 [mostly asymptomatic], then I2 if notable symptoms, then I3 if ideally hospitalized) that connect via a lattice of fluxes to two “resistant” (R) states. Here Nrec (“not recovered”) represents a placeholder for better tying emerging COVID-19 medical research findings with those from epidemiology.

**Methods:** Borrowing from fuzzy logic, a given model represents a “Universe of Discourse” (UoD) that is based on assumptions. Nonlinear flux rates are implemented using the classic Hill function, widely used in the biochemical and pharmaceutical fields and intuitive for inclusion within differential equations. There is support for “encounter” input events that modulate ongoing E (exposures) fluxes via S↔I1 and other I_1/2/3_ encounters, partitioned into a “social/group” (*u*_*SG*_*(t)*) behavioral subgroup (e.g., ideally informed by evolving science best-practices), and a smaller *u*_*TB*_*(t)* subgroup with added “spreader” lifestyle and event support. In addition to signal and flux trajectories (e.g., plotted over 300 days), key cumulative output metrics include fluxes such as I3→D deaths, I2→I3 hospital admittances, I1→I2 related to “cases” and R1+R2 resistant. The code, currently available as a well-commented Matlab Live Script file, uses a common modeling framework developed for a portfolio of other physiological models that tie to a planned textbook; an interactive web-based version will follow.

**Results:** Default population results are provided for the USA as a whole, three states in which this author has lived (Arizona, Wisconsin, Oregon), and several special hypothetical cases of idealized UoDs (e.g., nursing home; healthy lower-risk mostly on I1→R1 path to evaluate reinfection possibilities). Often known events were included (e.g., pulses for holiday weekends; Trump/governor-inspired summer outbreak in Arizona). Runs were mildly tuned by the author, in two stages: ***i)*** mild model-tuning (e.g., for risk demographics such as obesity), then ***ii)*** iterative input tuning to obtain similar overall March-thru-November curve shapes and appropriate cumulative numbers (recognizing limitations of data like “cases”). Predictions are consistent deaths, and CDC estimates of actual cases and immunity (e.g., antibodies). Results could be further refined by groups with more resources (human, data access, computational). It is hoped that its structure and causal predictions might prove helpful to policymakers, medical professionals, and “on the ground” managers of science-based interventions.

**Discussion and Future Directions:** These include: ***i)*** sensitivity of the model to parameters; ***ii)*** possible next steps for this SEI_3_R_2_S-Nrec framework such as dynamic sub-models to better address compartment-specific forms of population diversity (e.g., for E [host-parasite biophysics], I’s [infection diversity], and/or R’s [immune diversity]); ***iii)*** model’s potential utility as a framework for applying optimal/feedback control engineering to help manage the ongoing pandemic response in the context of competing subcriteria and emerging new tools (e.g., more timely testing, vaccines); and ***iv)*** ways in which the Nrec medical submodel could be expanded to provide refined estimates of the types of tissue damage, impairments and dysfunction that are known byproducts of the COVID-19 disease process, including as a function of existing comorbidities.

## 1 Introduction

### 1.1 Need for Dynamic Causal Models that Better tie Epidemiology with Medical Physiology

Our collective knowledge of SARS-CoV-2 as a virus from biophysical and bioprocess perspectives, and of COVID-19 as a disease from epidemiological and clinical perspectives, continues to rapidly evolve. Yet connections between such emerging knowledge domains tends to be heuristic. The usual tool, a pillar in epidemiology and often clinical medicine, is to use advanced descriptive statistical approaches (e.g., regression, covariates) to search for correlations, then try to use such insights to inform public health policy and medical intervention strategies. Such statistical methods, while powerful and capable of being extended to time series fitting, tend to be curve-fitting “trend analysis” tools that do not attempt to capture the underlying naturally-occurring causal dynamic processes that evolve over time. What may be happening “under the hood” can only be inferred.

This motivates a need for dynamic models based on differential equations. While such equations have been a pillar behind modern societal transformation for over a century, the epidemiology and medical fields tend to have a mathematical foundation based in statistics, and typically restrict use of ordinary differential equations (ODEs) to relatively simple first-order linear ODE’s such as those used to conceptualize “exponential growth” and “exponential decay” behavior. This statistical orientation reflects a recognition that physical equations cannot easily capture the behavior of what are frequently called Complex Adaptive Systems (CAS). But dynamic models can serve as a helpful tool, and the one offered here may provide one part of a problem-solving toolbox. It can add value by helping to refine one’s knowledge and discourse through the explicit act of model-building, to establish causal relationships between dynamic phenomena, and to estimate “under the hood” phenomena.

Relevant dynamic models mostly fall under two key categories: patho-physiological and epidemiological models. Dynamic physiological models take many forms, including “bicausal lumped parameter energetic” (electrical, mechanical, biomechanical roots), input-output (systems identification, electrical roots), and compartmental (biochemical, pharmaceutical, population biology roots). Examples of biophysical models applied to COVID-19 include the study of material flow dynamics of the respiratory system, which by Spring 2020 had helped establish airborne, aerosol transmission and the overriding importance of mask use (1,2). Traditional models of epidemiology date back to a sequence of papers by Kermack and McKendrick starting in 1927 (3). Such approaches fall under the framework of compartmental models, which have been widely used in physiology and biochemistry for a rich diversity of applications and become the natural tool for building a “mixed” model.

#### Epidemiological Compartmental Submodel

Each compartment is assumed to be well-mixed, with its “state” captured by a single variable representing an “amount” of something. This is most commonly a material species, but a subpopulation of a species (as in epidemiology) also works well. Between compartments are fluxes. By convention the outflux from one compartment equals the influx to the downstream compartment (any “loss” is a separate outflux to the “environment”). Borrowing from fuzzy systems theory (4), here we use the concept of a “Universe of Discourse” (UoD) to define an “ecosystem” consisting of idealized flow of subpopulations.

In traditional epidemiological modeling (5-13) the UoD is often defined by an abbreviation – SIR, SIS, SEIR, SEIRS – where here compartment S represents the susceptible subpopulation, E is exposed, I is infected (or infective), and R is variously resistant, recovered, or removed (i.e., immune to new infection). In this UoD of idealized well-mixed compartments, individuals reside within one compartment even though degrees of infection and resistance (and recovery) may coexist within a body as viral distribution, immune response and recovery processes evolve internally. Further, compartments lack diversity (a challenge, since a subpopulation within a compartment may have many attributes, and no one metric may be able to capture “heterogeneity”).

SEIRS-type models, often in simple forms, have been used extensively to illustrate epidemiological principles. In most cases, the flux rates have been assumed to be constants, often with one exception for the S→E flux: a modulated distribution “force of infection” or “immunity factor” called λ (14-16) or multiplicative flux modulator called β in the literature. More sophisticated models use statistics and covariates to obtain λ or β (e.g., 11) or include multi-SEIR models for partitioning age, spatial location, etc. (9,12,17,18). Principles that have emerged from such modeling efforts include: formalizing the Reproduction Ratio (*R*_*o*,_ secondary cases/case) as a function of rates for various model formulations that can include additions such as birth rates (5-8), illustrating advantageous properties of successful pathogen mutation (5-7), identifying conditions for stability and steady-state behavior (5-8), identifying conditions for a “herd immunity threshold” (HIT) (14-16), and accommodating seasonal fluctuations (11,13). As subpopulations such as S change, often an “effective” or “instantaneous” reproductive ratio *R*_*e*_ (12-16,17), *R*_*i*_ (19) are calculated over time for a deterministic or stochastic model.

A challenge with such models has been their use for everyday phenomena, such as spikes related to events (11). Our alternative approach is motivated by extending a respected SEIR hybrid model (11) which used two I states in series and a *β*-drive, while partitioning for severity (8,17). It is based on a “systems engineering” perspective, as introduced in **Figure 1** and described in Methods **Sections 4.1-4.4**, with a I-R lattice and β*(t)* expanded.

**Figure 1:**
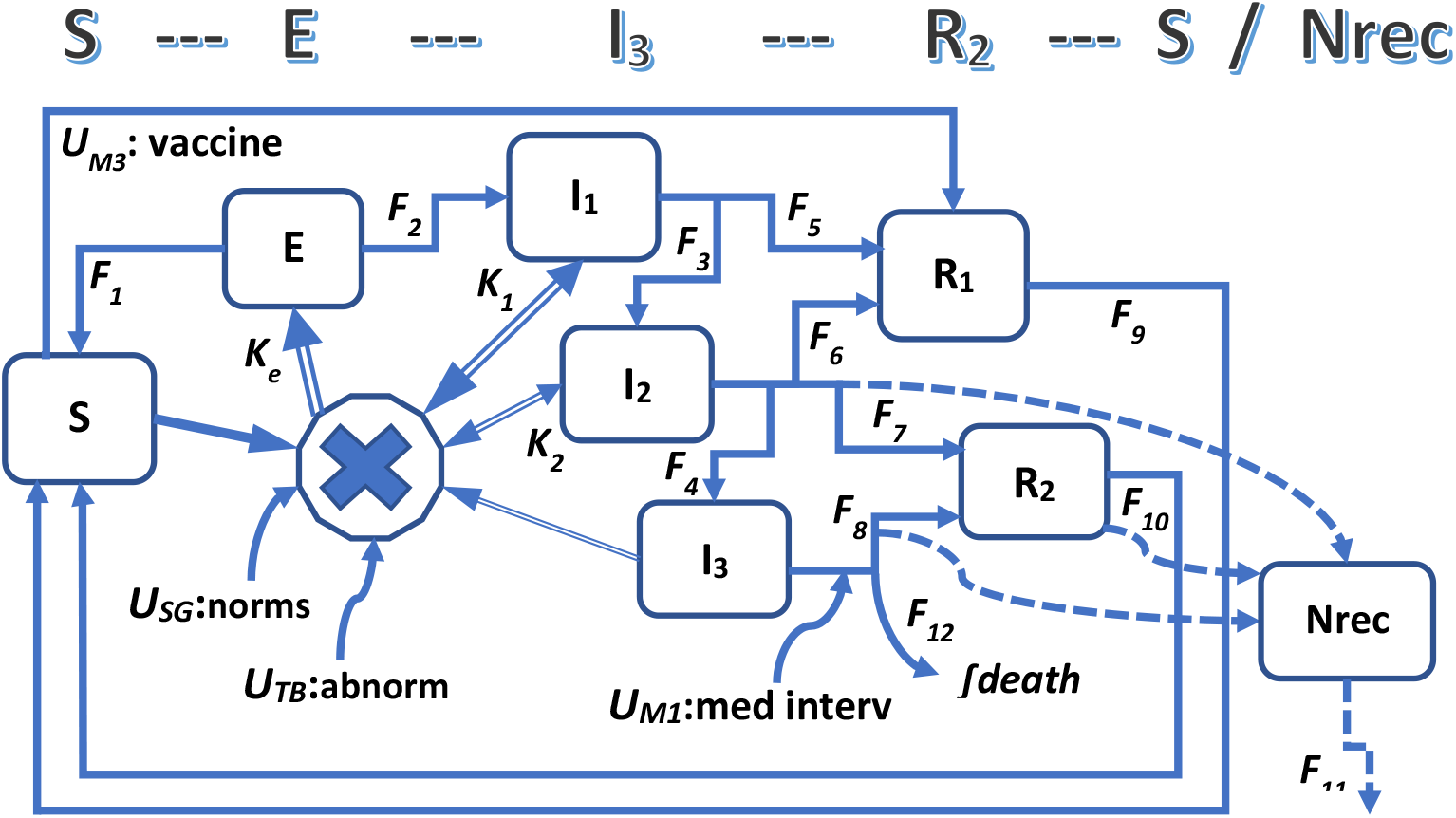
Conceptual diagram of the model. The SEI_3_R_2_S epidemiological submodel network consists of 7 subpopulation signals: S, E, I1, I2, I3, R1, R2. These are represented mathematically by normalized, dynamic states: *x*_*S*_, *x*_*E*_, *x*_*I1*_, *x*_*I2*_, *x*_*I3*_, *x*_*R1*_, *x*_*R2*_. At the start of a simulation (*t=0*), *∑x*_*i*_ *= 1*.*0*, i.e., by design, the summed population is 1.0. Arrows represent fluxes, with units of population/time. The large “X” represents multiplicative “encounters” between two signals that are modulated by an input trajectory representing subpopulations that mostly followed social directives (*u*_*SG*_) that are hopefully science-informed, and another chooses not to follow such directives (*u*_*TB*_). While S↔I1 interactions dominate actual exposures and model behavior, the 2-way arrows recognize that other encounters are possible (S↔I2, S↔I3, I1↔I1, I1↔I2, I1↔I3, I2↔I2, I2↔I3, I3↔I3). Compartmental signal S (susceptible) is normally by far the largest in size, e.g., starting at *t=0* over 0.999 (99.9%) and rarely dropping below 0.7 (70%). For the 3 stages of infection signals (I1, I2, I3), I1 (mostly asymptomatic) has the highest range and I3 (severe, ideally hospitalized) the lowest, but collectively they rarely reach over 1% of the population. The I-R lattice of 6 fluxes, a key contribution of this work, dramatically influences dynamic behavior. Resistant signal R1 trajectories are much larger than R2, and for some simulations (minus vaccine) these can sum to over 10% of the population within 6 months. Medical interventions *U*_*M1*_ and *U*_*M2*_ modulate fluxes, with *U*_*M1*_ lowering deaths and increasing the I3→R2 flux, and *U*_*M2*_ (not shown for clarity) enhancing the I2→R1 flux. Intervention *U*_*M3*_ represents emerging vaccines, and while shown as a S→R1 flux, is by default a weighted distribution to both R1 and R2, to be updated as we learn more. In dashed, Nrec (**N**ot **Rec**overed) is the placeholder for the medically-oriented “not recovered” state.

One feature of the SEIRS submodel, as used here, is that fluxes conserve total population N. Here states are normalized (“outputs” can be mappings, such as to “per 100,000”), and thus **S + E + I1 + I2 + I3 + R1 + R2 = N – D**, where here I1 is mostly asymptomatic, I2 is symptomatic, I3 is more severe, and R1 returns faster than R2 to S.

More formally, with N=1 and states *x*_*i*_*(t)* that evolve over time:

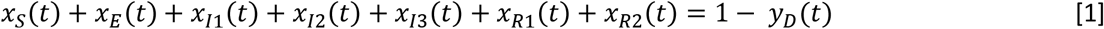

where this should naturally be satisfied at every time step in a simulation, and where *y*_*D*_*(t)* is an output (sum of a “death” flux) that accumulates over time. Thus, for this default UoD, the overall population does decrease.

Within the **Methods** section, the 7 state equations are provided in the **Section 4.1, Equation 3**, types of fluxes in **Section 4.2**, design strategies for science-informed ballpark estimates of flux rates in **Section 4.3**, and the rationale for each of signals E, I1, I2, I3, R1, and R2 in **Section 4.4**.

#### Medical Recovery Model

In SEIRS models, individuals eventually transit (hopefully) from I (infected) to R (resistant/recovered) states, then eventually back to S. It is important to recognize that temporal dynamics of this R state cannot be expected to correlate with **medical** recovery. As a physiological modeler, this distinction was initially difficult for this author, who “thinks” in terms of recovery (e.g., in medical rehabilitation this often involves tissue remodeling, organ function). Many of the observed and well-documented COVID-19 health challenges can continue in patients (and discharged patients) well after they would be classified as “resistant” to new infection (i.e., transitioned to R). These include tissue damage (e.g., within lungs, cardiac muscle, kidneys) and diminished organ function (e.g., lung capacity, sense of taste/smell, cognitive). For example, consider this recent quote of a proposed COVID-19 intervention, where the word “recovery” has been bolded for emphasis:

> “Results … received remdesivir had a median **recovery** time of 10 days (95% confidence interval [CI], 9 to 11), as compared with 15 days (95% CI, 13 to 18) among those who received placebo … Conclusions … show that remdesivir was superior to placebo in shortening the time to **recovery** in adults who were hospitalized with Covid-19 …” (20)

While the content remains an open issue as not all studies agree, here the key is to notice that “recovery time” in days is used in the typical medical context. Here “recovery” should not be expected to translate to a “resistant” state, or to an “infection” state, even here for a drug targeting the immune system. This is one of manifold examples. Furthermore, these dynamic processes of R (as resistant) and recovery are neither in series (i.e., sequential in time) or parallel (operations that later converge back). While all of I and R and “recovery” may be different constructs, there are methods that enable epidemiology-medical co-existence within one overall model. So how do we couple these differing concepts within our UoD, and do so without violating **Equation 1**?

Here we offer the state Nrec (**N**ot **rec**overed) as a simple “placeholder” to illustrate how a clinician-oriented “medical” submodel can be coupled with a SEIRS epidemiological submodel, both co-existing within one UoD. We start by accepting the concept that an **individual** within a population can have **degrees of recovery**. Using fuzzy logic, a classic AI (artificial intelligence) tool, we define “**not recovered**” within an interval <0,1>, where a “0” implies full recovery, a “1” indicates death (of an individual within the population), and a degree of “partially not recovered” rests between these extremes. Ideally, Nrec includes both acute recovery and chronic health challenges, positive effects of medical interventions (e.g., 20-22), and possible accumulation with reinfection.

In the default model, Nrec is a nonlinear function of I3 and I2 (e.g., via outfluxes to R2 and to death), and mildly of R2. Nrec could also be **bi-directional**, and influence these (or other) states (e.g., challenging recovery slowing I3 to R2), but only through fluxes since the total SEIRS population constraint must be protected. Nrec thus represents, for now, the summed collective effects of many individuals. But this NREC distribution could naturally extend to specific types of functional impairment or dysfunction (e.g., subgroups based on evolving data), either as calculated outputs or dynamic states (the latter would expand the order of the overall model).

The current Nrec state equation is provided under **Methods, Section 2.1, Equation 4**.

### 1.2 Need for Dynamic Models that Connect Societal Behavior (including “Spikes”) to SEIRS Models

The SEIRS class of models have been driven at the S→E transition in many ways, for instance by a flux modulated by β (contact rate multiplied by probability of transmission per contact (11)). We will offer an alternative approach that borrows from the tradition of “systems and control” engineering, after reviewing alternatives.

A rich line of epidemiological research is based on developing collective social interaction **contact networks** that can facilitate viral transmission, often using “big data” analytics. Examples include use of mobility geolocation datasets (e.g., for metropolitan Portland area (23-24), larger regions (17,18,25,26)). These can be helpful, but also controversial (e.g., huge variation in estimates of degree of spread through events such as the Sturgus Motorcycle Rally, up to hundreds of thousands). Studies have also considered roles of modern transportation on world-wide infection spread via host translocation operating through emergent “hub-spoke” “small world” complexity models (27) where “hubs” can be brain regions/cities/airports/web-apps and “spokes” involve local spread that evolves in space and time (25,28). Some “metapopulation” models have even mapped geospatial data to the types of facilities where people gather and the contexts associated with such encounters (25), or estimated risks of local gatherings based in part on emerging serology data (29, https://covid19risk.biosci.gatech.edu/), or used machine learning/neurofuzzy tools (30). Such models and tools, while intriguing and of value, may be limited as they infer rather than directly document the form of **actual encounters** between people, and tend not to be able to address important contextual information such as mask use (but could help drive SEIRS models).

Still another body of work is based on combining complex connection networks and/or geolocation “spacetime” network-type models to SEIRS-type compartmental models, which can be challenging since the two UoD’s tend not to mesh well. Such approaches involve making assumptions on networks, using SEIR networks with partitioned subpopulations such as by age (12,17,18) or make heterogeneous connectivity distribution networks usually assumed to persistent over time (13,15), and have developed a strong theoretical foundation (12-15). A key feature is capturing **population heterogeneity assuming a unimodal distribution** with a single mean and then a spread, for instance, with examples of assumed variational distribution including normal (Gaussian), binomal, Poisson, a power law, or centered exponential decay. This leads to theoretical elegance, and such models have been useful for past targets such sexually-transmitted diseases. One nice simplification has involved combining two λ’s in series, one for societal and one for biological variance, each representing a “heterogeneity” distribution and assumed to be independent of each other (15). Such models tend to be best equipped for evaluating steady-state behavior, formulating various reproduction ratios, and estimating HIT. These have included low HIT estimates (16) determined in ways that this author found to be problematic, which helped motivative the need for this paper. At the other extreme are individual-based (or agent) models (31).

From this author’s perspective, there are 3 concerns, all related to features of this specific COVID-19 pandemic: ***i)*** both social or physiological **heterogeneity** are difficult to capture with one metric (e.g., social mobility is dwarfed by the now-recognized importance of personal mask use (1,2,32-38) and indoor gatherings (29,32), physiological by well-documented factors such as age and comorbidity (39-42), see also **Methods Sections 4.4**); ***ii)*** population distributions are likely at least **bimodal** (e.g., social political factors; risks of young/healthy versus older/comorbidities); and ***iii)*** social behavior (43) and physiological status is **not persistent** but rather changes over time (e.g., social as directives or risks change, physiological with isolation and less access to healthcare).

An alternative approach is illustrated by the highly-respected work of the Institute for Health Metrics and Evaluation (IHME) group, which uses a clever hybrid approach that augments extensive statistical analysis of covariates (e.g., deaths, socially-distancing mandates (SDM), mask use) to help tune a SEIR model to back-estimate β, which is subsequently used to provide projections into the future (frequently cited by the press), given certain alternative assumptions for non-pharmaceutical interventions (NPI) such as mask use and SDMs (11). Use of such covariates can narrow CAS behavior into a more tractable problem, one that (to this author) has scientifically evolved toward 2 overriding considerations that emphasize airborne transmission and personal choice: mask use plus limiting indoor gatherings, in the context of **containment metrics and directives** (CMDs).

To this author’s knowledge, past methods have not been combined through a “controls-like” dynamic interface (close are 23,43). The **proposed model structure** offers an approach that builds on: ***i)*** the IHME SEIR model extended to an **I-R lattice** and with a loop back to S (**Figure 1**), plus with β now viewed as an input that modulates fluxes, and ***ii)*** a partitioning of this “encounter-modulation” interface input as consisting of two signals that naturally represent the conflict in our current “bi-modal” world that is partitioned into two types of actors, as motivated below and developed in **Section 4.5**. While still idealized, the **“**encounters” concept serves as a useful way of conceptualizing and simplifying the many social CAS phenomena into the **essential features** of social interaction contact moments, whether associated with transportation, work, local services, school, home, etc. A useful analogy is of motor neurons serving as the “final common pathway” for expressing collective brain behavior through motor neuron pools leading to muscle action. Here in the **encounter-modulating input** *u(t)* is assumed to function as the “final common pathway” of the collective interactive encounters of virus-carrying “hosts” with other members of the population. Conveniently, “encounters” can also represent biophysical host-parasite interaction, as will be illustrated in **Section 4.4** for ways to view the E compartment.

Our UoD assumes the SEIRS “subsystem” is “closed” but that inputs can modulate flux rates, similar to how λ or β are often used, but more general. Input-modulated paths are very common in physiology (e.g., enzyme-modulated biochemical reactions; hormones such as insulin modulate material fluxes such as glucose into cells such as muscle). Similar to predator-prey and such biochemical reaction networks, we will we assume that our inputs modulate “encounters” between two “species” signal variables, at least one of which is an I (infected) subpopulation (the other is most notably S, but we will also support modulated I↔I encounters only at lower sensitivity and occurrence frequencies, see **Section 4.5, Equation 7**).

We break down this dynamic input as the sum of 2 idealized stereotypes that reflect and classify two types of responses to the pandemic: ***i)*** a majority that mostly complies with “common good” social/group CMDs (*u*_*SG*_) and hopefully science-based guidance (67% by default); and ***ii)*** an individualism-or tribe-first first minority that doesn’t tend to comply and can be viewed as tribal/bellicose (or Trump/Bolsonaro, in honor of representative advocates), *u*_*TB*_. Both include a deterministic (*-det*) and stochastic (*rn*) component (see also **Figure 2**):

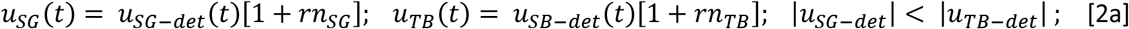

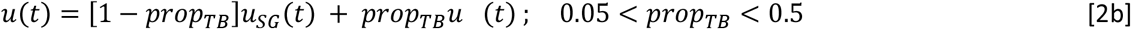

where we assume that for every moment in time, the normalized deterministic component of the input for “social guidance” behavior is less than that for “Trump-Bolsonaro” behavior. Whether *u*_*SG*_ or *u*_*TB*_ has a greater influence on the overall driving input *u(t)* depends on the proportion *prop*_*TB*_ between the two contributions, but usually if *prop*_*TB*_ is above about one-quarter (25%) we will see that *u*_*TB*_ tends to dominate results.

**Figure 2:**
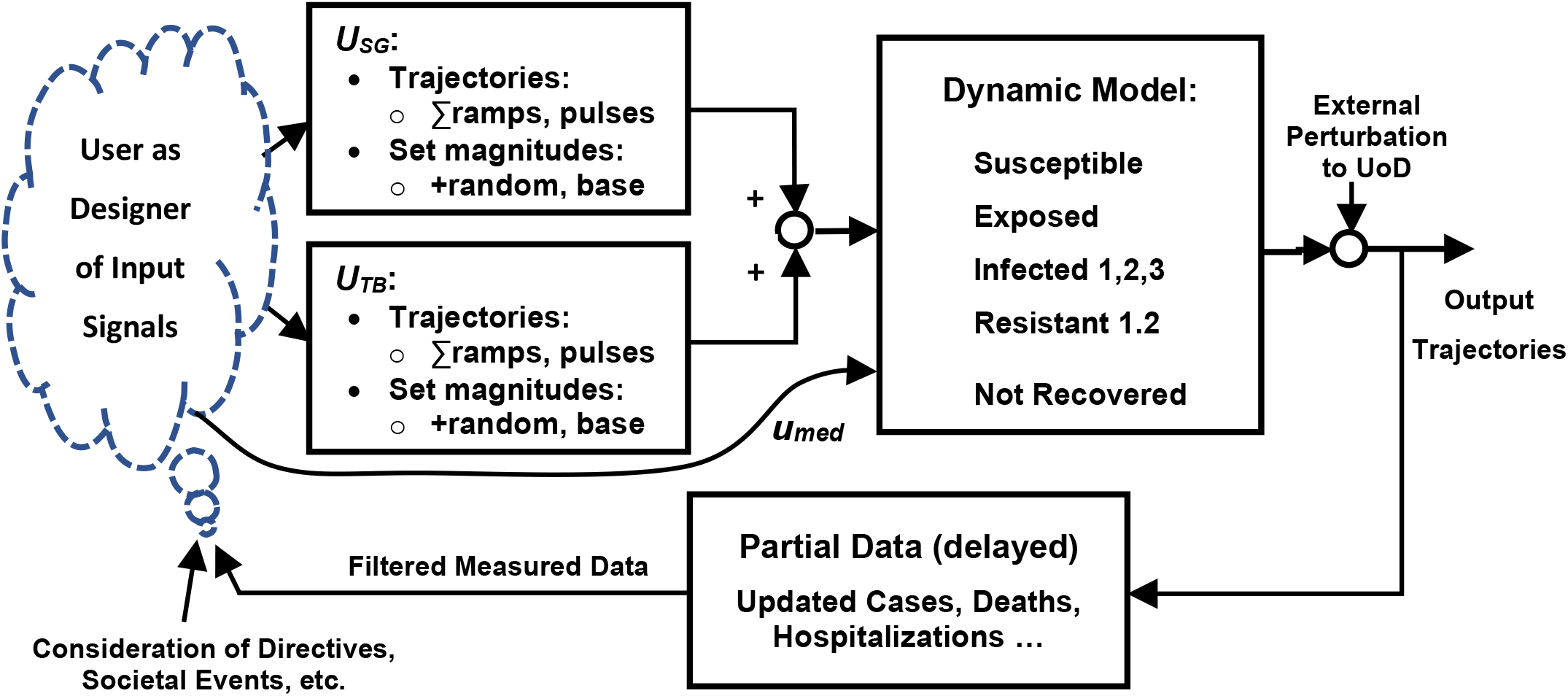
Design considerations for Forward Dynamic simulations. Given a model and a UoD, the user comes up with the input trajectories by synthesizing available information such as societal directives and “spreader” events plus, if available for all or part of the simulation time, pandemic data that is applicable to the UoD. The 8 states and the outputs (trajectories, cumulative metrics) then evolve over time. Since there is a random component augmenting the prescribed input trajectories (adds to deterministic component of signal), repeated simulation runs will not be identical, and while it is theoretically possible for dramatic changes between such runs for this nonlinear model, this has not been observed. The *u*_*med*_ interventions mostly operate on fluxes (e.g., lower death flux rate, some other fluxes). If the external perturbation is used, it would usually represent added I1 infection(s) (“importations”) to the UoD at a specified time (e.g., airport arrival(s)). While this heuristic, iterative tuning approach was used for all of the simulations presented here, advanced statistical and optimization tools are available in Matlab toolboxes, and use would involve straightforward coding adjustments (although optimization for many-month simulations may require considerable computational resources).

Formulation of the strategy for creation of input trajectories is provided in **Methods, Section 4.5**.

### 1.3 This Contribution in Context

A model is a tool, hopefully adding value through simulations that: ***i)*** provide insights in causal behavior (e.g., relations between input drives and output trajectories/cumulative metrics), ***ii)*** estimate internal phenomena that is difficult to measure (e.g., “hidden” infections I1), and ***iii)*** make helpful predictions of future behavior under certain assumed conditions (e.g., relative proportion *u*_*SG*_ versus *u*_*TB*_ subpopulations).

Key contributions include: ***i)*** turning the SEIRS model into a small lattice of connectivity, ***ii)*** framing the UoD and model structure using a “control engineering” perspective, and ***iii)*** offering an interactive model for users.

There are limitations to this framework, and the original plan of this author was a more sophisticated and “AI-based” model. But it is best to start simpler, and the approach that evolved was found to be strikingly non-trivial, with rich results that tied to COVID-19 data (e.g., cases, deaths), and worthy of serving as a foundation for improvements. Suggestions for possible future directions are offered in **Discussion Sections 3.3 to 3.6**.

## 2 Results

For Methods section, see **4 Methods** (starts on page 18).

### 2.1 Overview of Model Simulations

Most of the simulation results presented for this paper run for a time period of 300 days, from March 1 through most of December. As a novel model, initial “starter” model parameter values were best estimates based on a synthesis of biophysical and biomedical processes associated with each rate (see **Section 4.4**), then heuristically adjusted. Here are the default values, in units of *x* for *k*_*hal*f_ and /day for rates, extracted from Matlab code:

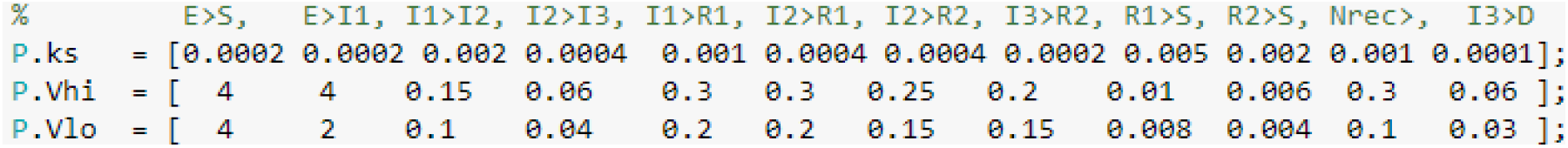

Notice that in all cases parameters have one significant digit, or in a few cases, a second digit that is a 5, i.e., the halfway point. This is on purpose, to illustrate that the primary contribution here is the model structural design. Some could argue that E>I1 (helps give “latency”) and R1>S (time “resistant”) could be lower, and this can work. These values were sufficient to provide reasonable estimates for not just USA simulations but also the 3 other states considered plus some special cases, given reasonable assumptions to produce driving inputs. What is offered here is a rather robust platform for future tuning by other groups with access to more resources.

Methods **Sections 4.3-4.4** provides suggestions for adjustments based on demographic data. For instance, Oregon has a well-documented lower obesity rate than the USA as a whole, and Wisconsin slightly higher – these mildly affected some flux rate parameters (by 5-10%) and thus fluxes between I and R compartments (thus affecting downstream distributions), which in turn has some effect on fluxes to Nrec.

The default code, for all cases presented here, is available as Matlab Live Script file ch2_covid19_JWv2. Unlike other models this author has designed, systematic sensitivity parametric analysis is not supported for this script code version but will be available on a subsequent interactive web version. Of note is that model behavior has been found to be very sensitive to many model parameters, beyond those associated with the E→I1 flux that would be expected based on past studies. Importantly, in contrast to traditional SEIRS serial cascade models, rate parameter modulation within the I-R lattice can lead to strong cross-coupling sensitivity effects.

Most simulations of larger populations started with a very small subset of the population expressing symptoms (i.e., state I2), e.g., an initial state of ***x***_***o***_ = [0.9999 0.00005 0.00004 0.00001 0.0 0.0 0.0 0.0], where the first 4 values represent S, E, I1, I2. The I1 starter value is more important than E, but both express the reality of an interacting population with a small “hidden” I1 at time *t=0*.*0* prior to detection after some I1→I2. From then on, inputs simply modulate fluxes, and the simulation evolves. For intuition, scaling is such that input drivers ***u***_***SG***_ and *u*_*TB*_ input drives is such that “1.0” is near-normal, modulated by time of day (lower at night) and quasi-random noise pulses that augment the deterministic portion of the signal (these represent the fluctuations of collective daily life). Thus, the model is stochastic, and runs with identical input designs may differ slightly.

### 2.2 Systems Analysis: Sensitivity to Step and “Pulse” Response Magnitudes

There is a long history of using SIS, SIR, SEIR and SEIRS models to illustrate principles such as stability, steady-state behavior and reproduction ratios (*R*_*i*_’s). **Section 4.5** and **Equ. 7** showed that for this model, with inherently nonlinear rates and fluctuating daily inputs (e.g., lower at nighttime), it is not that simple. But the usual insights still hold. *R*_*i*_’s are still dominated by the relative magnitude of the input-modulated influx to I1 from E versus the 2 outfluxes of I1 (since I1→R1 >> I1→I2, I1→R1 is more dominant). The model was “designed” such that *u*_*SD*_ and *u*_*TB*_ ramps 0.8-1.0 will yield “ballpark” results (see **Figure 3**). Notice that there is a month or so of (exponential) latency. Results are very sensitive to the step value within the critical 0.7-1.2 range. **Table 1** summarizes key metrics. As perspective, for the USA runs, *u*_*SD*_ averaged below 0.8 (started at 1.0) and *u*_*TB*_ about 0.9 (started 1.2), clearly within sensitive “deaths” range, with peaks even worse. Notice huge S and R_tot_ ranges (sum to ∼99%). Clearly getting *u(t)* ≈ 0.7 is good for CMDs, and above 0.9 is problematic.

**Table 1:** Selected summary of key metrics for systems analysis “step” runs. Some are at time slices (180, 270), some cumulative to 270 days.

| Step $u_{SG}$ & $u_{TB}$ | 0.7 | 0.8 | 0.9 | 1.0 | 1.1 | 1.2 | 1.3 |
| --- | --- | --- | --- | --- | --- | --- | --- |
| Deaths by 270 | 0.003 | 0.15 | 0.27 | 0.40 | 0.55 | 0.63 | 0.74 |
| S@270 (%) | 99.6 | 87.3 | 86.1 | 78.5 | 70.2 | 67.0 | 65.7 |
| $I_{tot}@270$ (%) | 0.019 | 0.18 | 0.27 | 0.69 | 0.64 | 0.39 | 0.29 |
| $R_{tot}@180$ (%) | 0.20 | 7.5 | 19.2 | 20.0 | 20.8 | 22.3 | 25.9 |
| $R_{tot}@270$ (%) | 0.36 | 12.3 | 13.4 | 20.3 | 28.8 | 32.0 | 33.2 |
| Nrec@270 | 0.01 | 0.12 | 0.15 | 0.22 | 0.27 | 0.28 | 0.29 |

**Figure 3:**
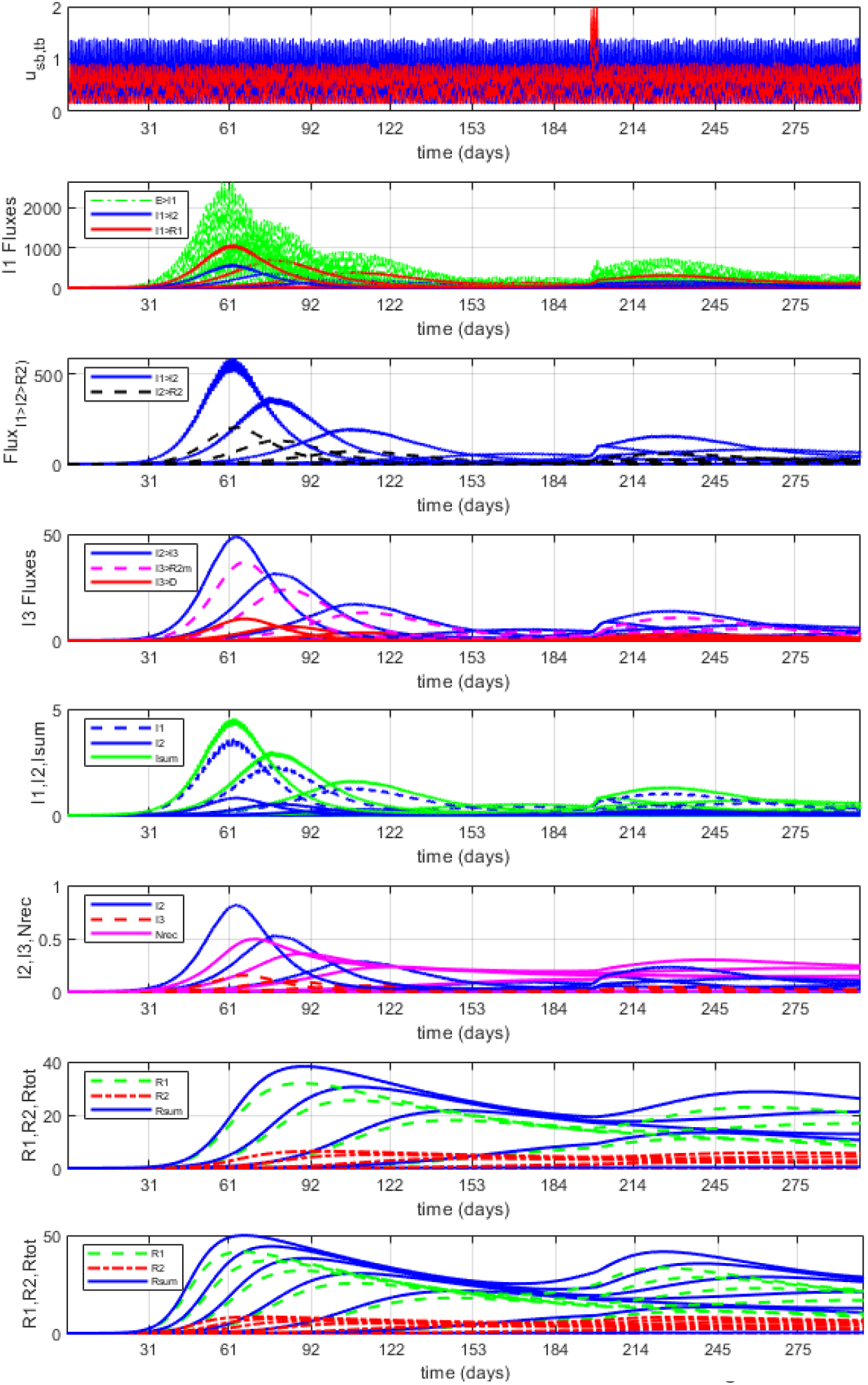
Overplotted Step+Pulse Responses for *u*_*BG*_= *u*_*TB*_= {0.7 0.8 0.9 1.0 1.1}. Run is for 300 days (starting March 2020), initiated with very small I2 and I1 levels such that 99.9% starts in state S (as also used to start USA/States runs). Here instead of tuning ramps and pulses (e.g., see, **Figure 2**), “ramps” remain flat at the step level. There is then a single 3-day pulse, using the 4th of July weekend. As this is a multi-input model, we let medical interventions *uM1* = *uM2* = 1.2, a moderate value. The run for 0.7 can barely be seen, and runs for 1.2 and 1.3 are not shown because they dominate the curve ranges, except for the very-bottom R curves to illustrate the nonlinear behavior trends. Notice that even for higher step values, it takes a month of “stealth” time to show up on this scale, since the initial condition (tiny I1, I2) was so low. Also notice that exponential growth does not continue, and the system is **naturally stable**. The highest pulse-responses for fluxes shown near 200 days are for 0.8 and 0.9, as they have more S at this time. But more important are the trajectories and steady-state signals for the I’s and R’s, which after peaking “drift” towards a quasi-steady state. The drift is gradual because of the I2→S flux rate (looping back) is so slow. The death fluxes (4^th^ plot, red) are hard to see except for the 1.3 and 1.2 steps, but cumulative data is reported in **Table 1** that illustrates that “**flattening the curve**” is effective. Here infections never reach 1% until the 1.2 step drive (not shown), and tend to take the form of (delayed) “impulse” or “pulse” responses. The most functionally relevant may be the R values, which are reported in **Table 1** for t=180 and 270 days. Notice that by step 1.0, over 20% of population is in R (resistant) state. Fluxes (rows 2-4) are of less of a focus here, but notice fluxes get smoother downstream, and that with higher steps, signals rise sooner and have higher maxima. Finally, the Nrec placeholder also rises with step level (hard to see, see also **Table 1**).

### 2.3 Many Reasonable Input Streams Lead to Elimination COVID-19 Within 2 Months (Given Ideal UoD)

**Figure 4** provides an example of how “easily” the SARS-CoV-2 parasite can be irradicated with an idealized UoD in a cohesive society. Here this author, after tuning for the USA that included a large March-April “first wave” in parts of the USA (**Section 2.4**), accidently “killed off” the virus in a first iteration of reducing *u*_*SB*_ and *u*_*TB*_ slope drives, here for Arizona. This can happen if *u*_*SB*_ is responsive to an outbreak (e.g., below 0.8 during a time window) while *u*_*TB*_ is also relatively low, there is moderate population noise, and few spreader events. While similar in concept to the concept of *R*_*e*_ or *R*_*i*_ (i.e., reproduction ratios that update, see **Equ. 8**), this is a broader principle since it illustrates the effects of behavior due to dynamic input drives, and history-dependence.

**Figure 4:**
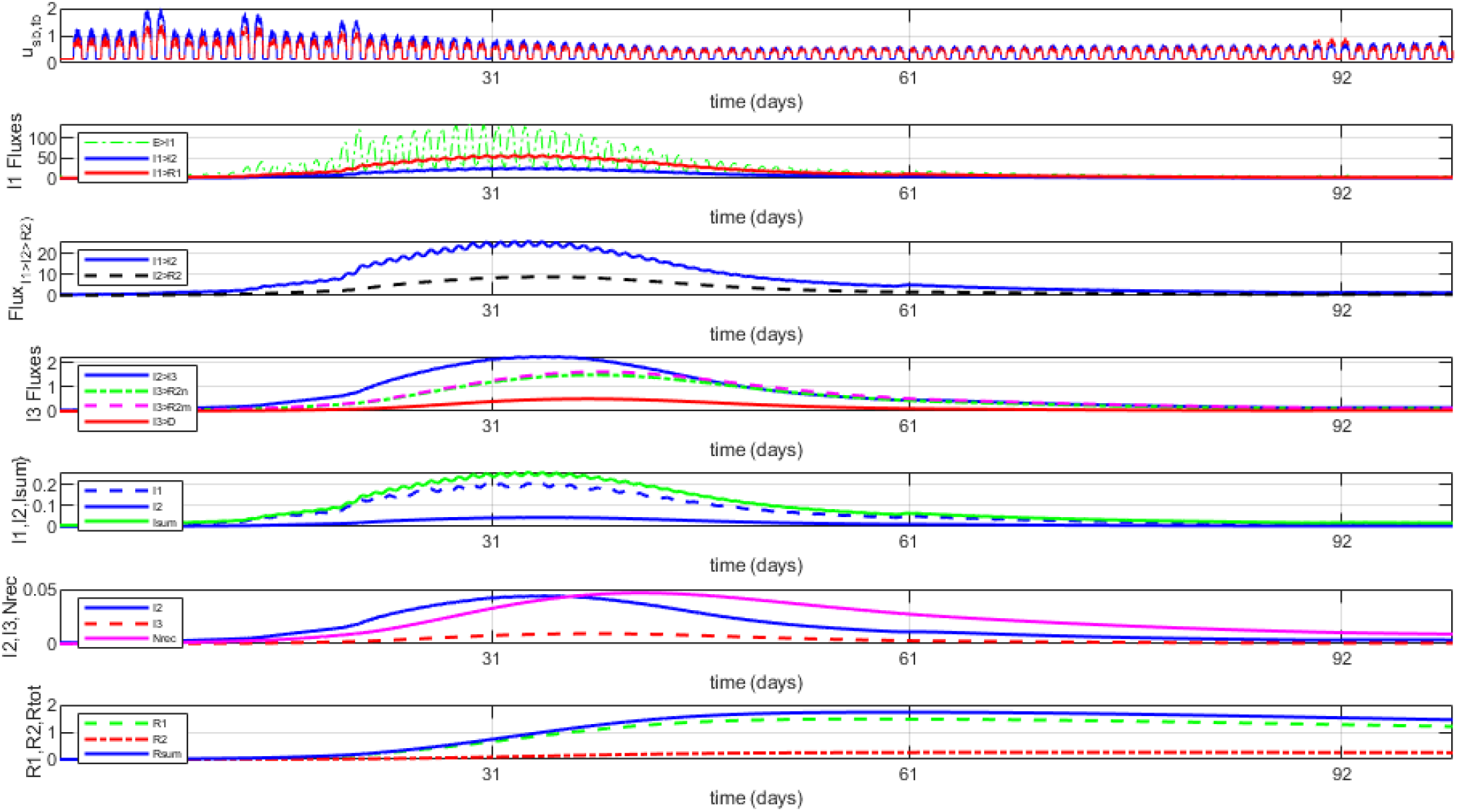
Example of 100-day March-to-early-June run where SARS-CoV-19 disease diminished. Starting from the USA default simulation (**Section 2.3, Figure 5**), a first attempt by this author at reducing slopes for *u*_*SG*_ and *u*_*TB*_ for use for Arizona (which was initially moderate) “accidently” killed off SARS-CoV-2. A new I1 “importation” event at 60 days and a Memorial Day weekend pulse at 89-91 are also well-managed.

The simulations of **Figure 4** run for only 100 days in part to help illustrate that there is day-night fluctuation (lower encounter opportunities during night), plus two related concepts: ***i)*** an added external I1 “bolus” representing an “importation” at 60 days (e.g., from airport, this also implies a very small addition to the overall population) had minimal impact (small “impulse response” that died out) because of CMDs in place and I1 being already low, and thus a spreader event did not emerge; and ***ii)*** a “normal” Memorial Day weekend “event” had only a minimal effect (the minor increase might need to be managed, but here the existing CMDs worked).

Such simulations give predictions that are similar to what has occurred in places that aggressively managed new infections, like China, New Zealand and Nova Scotia. Of course, it is difficult for society to be so cohesive (e.g., difficult to discourage *u*_*TB*_ pulse “spreader” events and keep signaling fluctuations to manageable levels).

Alternatively, there are also time windows where there is **high sensitivity to rapid growth**, especially when **I1 is growing (mostly undetected) relative to I2**. One example involves the quick decision by the NBA to suspend all basketball games starting on March 12, which was soon followed by the NCAA (no March madness), NHL, etc. This occurred just before a sensitive time window - large *u*_*TB*_ pulses in mid-to-late March (e.g., imagine full arenas, before high mask use) predict horrific results (both I3, deaths). Of note is that even in high-sensitivity time windows, “exponential growth” will gradually subside and change concavity, with I’s peaking then drop. For lower input drives, the classic “S”-shape will naturally reach a steady-state “capacity” signal distribution.

### 2.4 USA Default “Starter” Run

#### Overall USA input trajectory design

The input trajectories for the USA simulation were cautious, since: ***i)*** initial outbreaks were regional; ***ii)*** there was a disorganized federal response; ***iii)*** policies of various states differed dramatically; and ***iv)*** containment directives for SARS-CoV-2 started out as suboptimal (while there were severe “stay at home” shutdowns in many regions, evolving guidance was not yet based on aerosol transmission, where mask use and limiting indoor gatherings become higher priorities than, say, surface disinfectant cleaning).

See **Discussion Section 3.2**, including **Tables 2-3** for additional discussion on strategy and synthesis.

**Table 2:** Example of insights that can be extracted from signals, here from default simulations.

|  |
| --- |
| **Table 2:** Example of insights that can be extracted from signals, here from default simulations. |
| *S signal:* Roughly 88% of the USA population is susceptible as of late November (~4% of these for a second time). Even in Wisconsin with its surge over 75% of the population remains susceptible, and in Oregon it is still about 93%. |
| *Cumulative S→I1 flux:* Results estimate that over 25% of the USA population has been infected with the COVID-19 disease, and with assumed R1→S flux, over 1% of these having been reinfected (likely without their knowledge, as most of these would have transitioned through the low/no symptom I1 signal to the faster R1 dynamics). |
| *Current I1+I2+I3 signals:* November will end with about 1% of the entire USA population being currently infected. This is growing rapidly and could near 2% by late December. It had already reached 2% in Wisconsin by late November. |
| *I1→I2 flux:* Nearly 4% of the USA population has expressed symptoms (reached I2), higher than the 3% of documented cases (which in addition to I2 with symptoms, includes some I1 tested through contact tracing, etc.). This suggests that the USA case count of 14 million is way low, as is commonly assumed (e.g., CDC suggests a factor of 3, WHO). |
| *Hospitalizations:* If I3 plus a fraction of R2 (i.e., resistant but still recovering) represents people who should be hospitalized, hospitalizations have had 2 past peaks (as expected) and are now rapidly rising. |
| *D (deaths):* The only way to get the USA death curve to be higher in March-April and then lower is to assume that medical interventions have improved as we've come to understand how to better manage the disease, plus assume a nonlinear flux rate that is higher when I3 is high (e.g., hospital capacity reached, perhaps fewer resources/patient). |
| *Cyclic reinfections:* The model predicts that a subset of the USA population, especially a subset of people with inherent $u_{TB}$ behavior who naturally flow through the milder I1-R1 path, could easily reinfect members of the community twice per year. As currently tuned, the model suggests that thousands already have done so (**Figures 5-7**). |
| *Eliminating SARS-CoV-2/COVID-19 within a UoD:* For idealized UoD without external influences, it is embarrassingly "easy" to eliminate the virus within 2 months when a strong majority of the population follows $u_{SG}$ CMDs that are science-based and I1 drops low enough. The "no symptom" I1 is what matters, illustrating the importance of continued expansion of testing. Very low I1 is the key to fully reopening society, and even new external I1 perturbations can be handled if identified and managed through mild (but timely) adjustments in inputs. |
| *Exponential growth time windows naturally subside:* For the "closed" 7-state subsystem, periods of exponential growth will gradually subside and "flatten" for signals even with sustained high drives, as this is naturally a stable system. |

#### U_*SG*_ design

The basic slope changes were mild, from a start of 1.0 to a low reaching 0.6 at 45 days (mid-April) then rising toward 0.7 starting around Memorial weekend, then lowering slightly late summer as some containment directives were added, then gradually raising Fall 2020 as some businesses and schools reopened (especially in South and Midwest). With flu season, a modest seasonal value was added to slopes. Then during November, many regions added CMDs to try to lower outbreaks, and two alternatives for *u*_*SG*_ slope are given.

#### U_TB_ design

For the assumed 33% of the population, the lowest slope target value was 0.8, with a mild drop late summer, and then mild increases plus a mild seasonal flu augmentation. There were added pulses for each of Memorial, 4^th^ of July, Labor Day weekends, around and on Election Day, Thanksgiving (2 alternatives), plus mild pulses every weekend starting with the summer after Memorial Day. No “regional” spreader events were included (e.g., Sturgis rally). Rather, the weekend pulses were used are intended to be representative of how transient “impulse” responses can set the stage for higher model sensitivity to subsequent events.

#### UMED-1,2 design

It is well documented that medical interventions for COVID-19 have improved, both for preventing deaths (e.g., 44,45) and administering therapeutics (e.g., 20,46). As such, the code supports slope changes that, when over 1.0, multiplicatively adjust fluxes to R’s and D (death). These were assumed to increase over time, starting at 1.1, especially during the Summer, and by December were assumed to be 1.8 for *u*_*M1*_, 1.5 for *u*_*M2*_. A separate simulation where values remain at 1.0 predicts that the cumulative number of USA deaths would increase by 84%, I3 time goes up 33% (slower to R2) and the Nrec (not recovered) curve is much higher.

#### Mild heuristic tuning

There were two heuristic “fitting” criteria: ***i)*** cumulative numbers for fluxes I1→I2 (∼cases), I1→I2 (∼hospitalizations), I3→D (∼deaths), but with {I1→I2 > “cases”} because of testing barriers; and ***ii)*** matching the general shape of the curves for fluxes and states. This was accomplished within a limited number of iterations, without trying to precisely fit the available curves, especially given that USA numbers over time needed to be taken in context (e.g., low reported “cases” for the first few months, as access to testing was limited). “Hospitalizations” is also an imperfect comparison, as some people avoid hospitals and hospitalization stays would include not only I3 but also some R2 that are “resistant” but not yet sufficiently recovered to be discharged. Deaths (assumed via I3 outflux) may also relate to so-called “cytokine storm” (excessive immune response) in individuals who might be classified as resistant (R2). Thus, being reasonably close (and somewhat higher than) these values was deemed sufficient since it met the aim of illustrating the capabilities of the model.

#### USA results highlights

The general shape of the simulated USA curves, especially for the I1→I2, I2→I3 and I3→D fluxes, and the I2 and I3 curves, are similar to available data, especially when one notes that early “case” data should be well below I1→I2 (low access to tests) and the latter “case” data would also identify some I1 (e.g., via contact tracing). Predicted cumulative 270-day values, from March through Thanksgiving, are provided in the **legend for Figure 5**. All are higher, with I2 about double the “cases” count (as expected given limited early testing) and I3 is somewhat above the “hospitalizations” count. Total R results hover around 10% from late July through early October, similar to CDC and other estimates from SARS-CoV-2 antibody sampling (e.g., 47).

**Figure 5:**
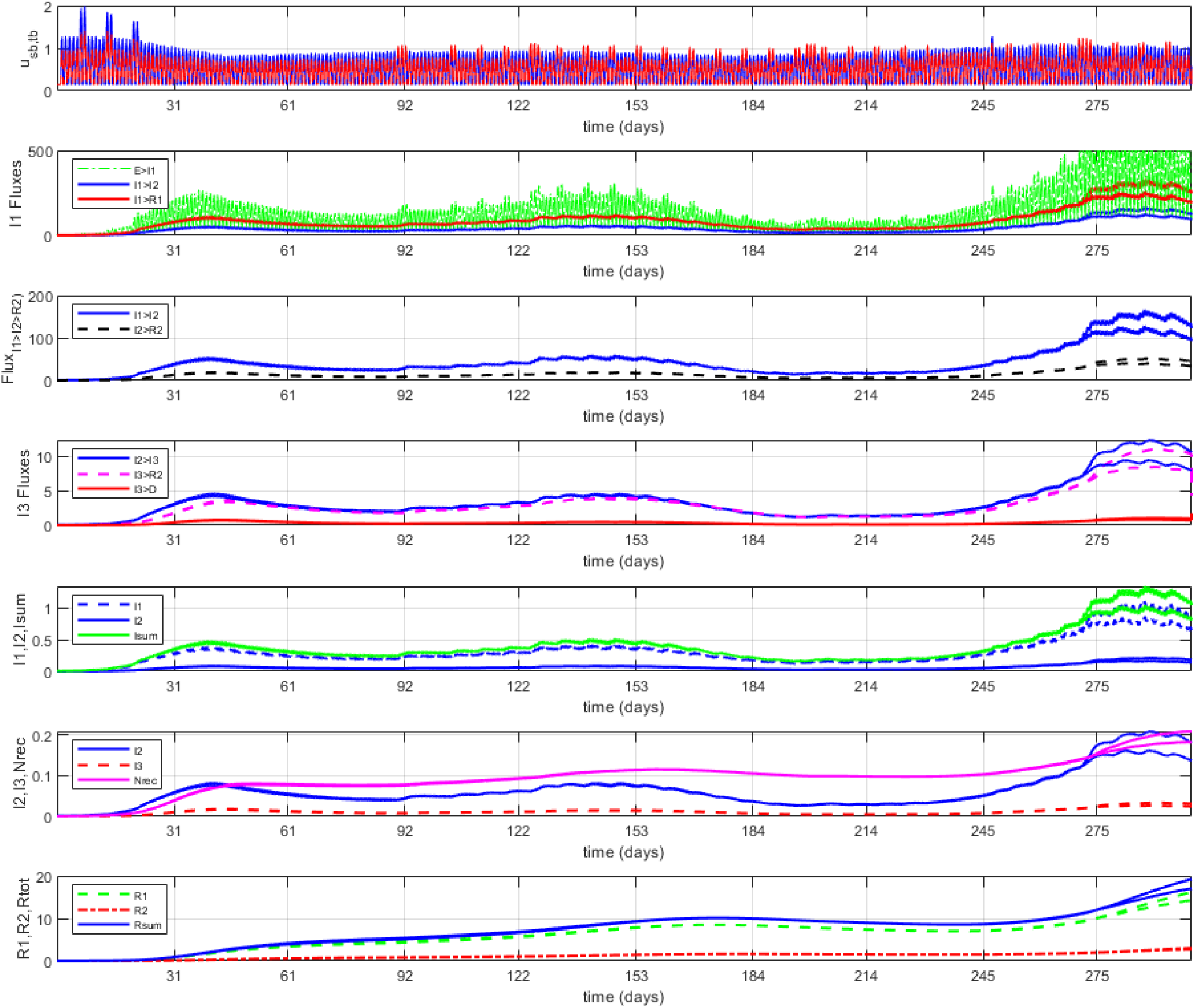
Selected plots (“Figure 1” from Matlab code) from the USA “starter” simulation (300 days). The top overplots the daily fluctuations in the *u*_*SG*_ (blue, solid) and *u*_*TB*_ (red, solid) inputs, which in turn drive the E→I1 flux. Plots for fluxes then follow, then signals. USA “cases” would most relate to I1→I2 flux (i.e., transition to notable symptoms) for early months (but lower) and then be closer to I1→I2 in later months as access to diagnostic testing (somewhat) improved. The flux I2→I3 (severe) ideally ties to hospital admissions, and I3 signal to hospitalizations (but both aimed to be slightly higher). Normalized states (bottom 3 curves) are multiplied by 100 and presented as a percent. Curves summing I and also R are provided. Starting just before Thanksgiving, 2 alternative predictions are provided, differing by whether new CMD adjustments are followed (lower of the 2 curves). Notice that total I’s may cross 1% of population by early December (after reaching about 0.5% at previous peaks), that R1 (mild R) is much higher than R2 signal, that D (deaths) has had 2 past peaks and is growing, and Nrec is always above D and is growing. Cumulative metrics were stopped at 270 days (i.e., through Thanksgiving), and values for these include, given per 100k (multiply by 3220 for total USA): **1)** for fluxes: 28k for E→I1, 8.7k for I1→I2, 0.7k for I2→I3, 22.4k for I1→R1, 22.4k for I1→R1, 0.116 for I3→D, and 16.3k for R1 & R2 →S (i.e., back to S, suggests >1k reinfected); **2)** for signals, as proportioned at 270 days: 88% for S, 0.58% for I1, 0.12% for I2, 0.019% for I3, 9.2% for R1, 1.9% for I2, and thus have about 0.72% of population infected for Thanksgiving (and growing) and over 11% for summed R’s (and rapidly growing).

#### General causal trend in weekly fluctuation

Weekend pulses can be used to illustrate transient dynamics in the model, where we tend to find a peak in I1→I2 (symptoms) about 3-6 days later (e.g., ∼Thursday) that stays high and only slowly returns, and thus a mild weekly fluctuation that can cause growth. Ironically, data from actual cases tend to also show a weekly fluctuation, which this author initially assumed was due to fewer PCR tests during weekends (fewer staff). But since there is now a 1-2 day shift in obtaining PCR results and since the USA “cases” days often peak around Friday, perhaps weekends really do tend to generate more spreader behavior.

### 2.5 Default Runs for 3 States: Arizona, Wisconsin, Oregon

The three states selected for simulation – Arizona, Wisconsin, Oregon – are places the author has lived, and knows well (two others, California and Maryland, made less sense since California is essentially like multiple states and Maryland is too interwoven with the DC and Virginia areas). All three have one major city that is not the capital, a clear capital with an embedded or nearby college town, and rural communities. All three also differed considerably in their approach to the pandemic, and some had well-documented opening-up policies and/or likely spreader events. Overplots of selected curves for the three states are provided in **Figure 6**. A more complete set of curves for the 3 states are provided in **Appendix 1, Figures 11-13**.

**Figure 6:**
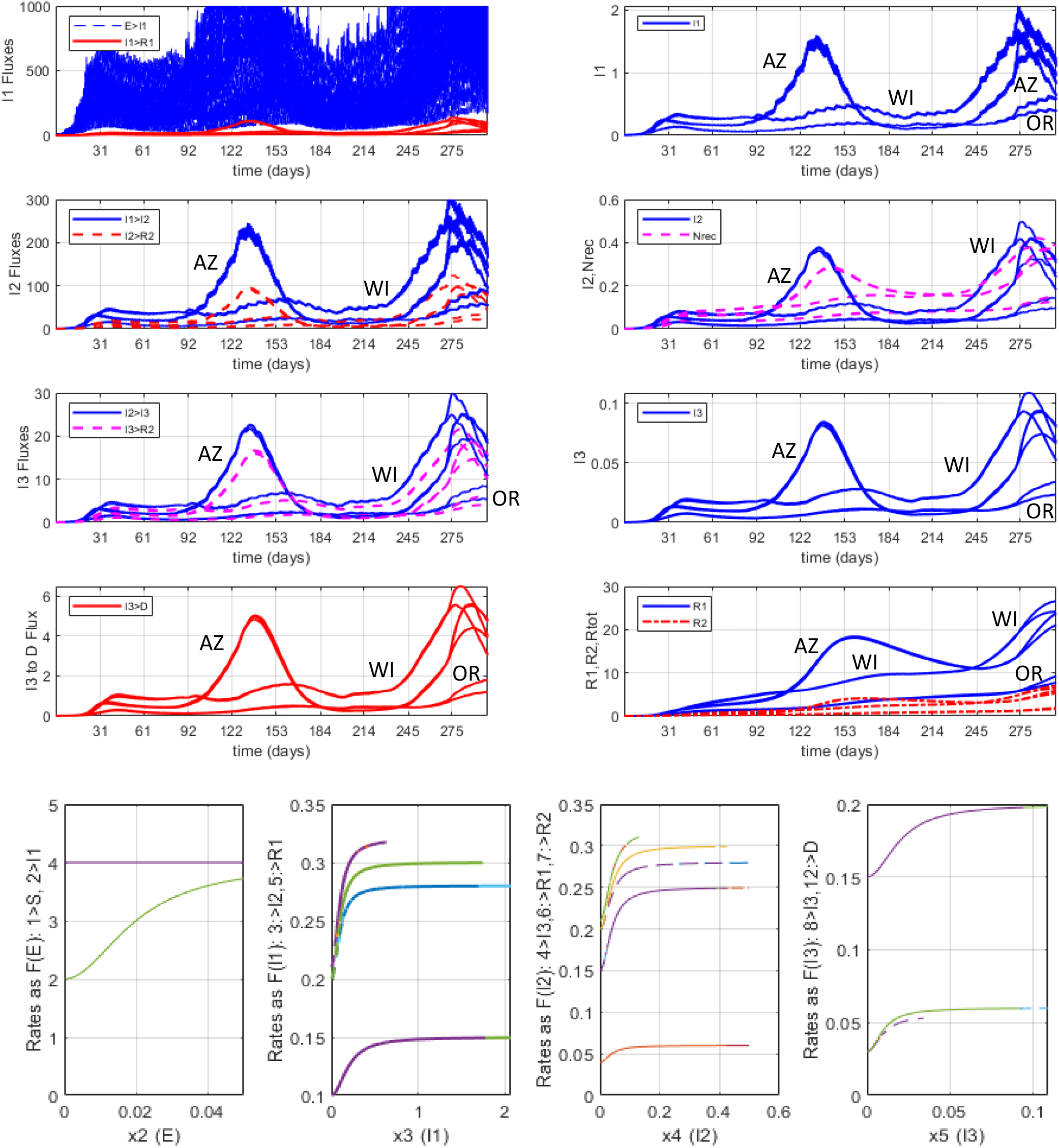
Selected overplots from “starter” simulation runs for states Arizona, Wisconsin and Oregon. ***Top Left:*** Key fluxes (E→I1,I1→I2, I2→I3, I1→R1, I2→R1, I3→R2, I3→D) are overplotted for the 3 states. ***Top Right***: Key signals (I2, I3, R1, R_tot_) are overplotted for the 3 states. See comments within code for some assumed dates for events, and see Appendix **Figures 11-13** for an isolated run for each state. For both here and in the Appendix, two alternatives are overplotted for Thanksgiving-to-December 25 predictions that differ in terms of CMD behavior and level of compliance of the *u*_*TB*_ subpopulation. Plots high for summer are for Arizona. ***Bottom***: Rates as a function of key signals (i.e., all are outflux rates), showing nonlinearity. Since these are overplotted, notice that some rates do not go as far on the x-axis, as the operating ranges were lower (this especially applies to Oregon). Also, certain rates for Wisconsin and Oregon differed slightly from the default, based on risk factors. Note: These are screen dumps from Matlab Figure 3 (top) and part of Figure 2 (bottom).

#### Arizona

For the first attempt at Arizona (Case 2 in code), which had a very low number of cases for March-April-May, this author lowered the *uSD* and *uTB* curves moderately and accidently killed off the virus (**Figure 4**). These were raised appropriately. The rising of Arizona curves during especially June (**Figure 4, Appendix Figure 11**) correlate with visits by President Trump where “opening up” was emphasized, and the sequence of choices by the governor before a subsequent visit (dates are documented in the Case 2 code comments). The inputs where adjusted by changing the *uSB* and *uTB* slopes to reflect the governor’s re-opening directives, and associate *uTB* pulses with events documented in the news. This was followed by a mostly community-driven containment process during late July and August which the CDC found effective (48). This summertime surge makes sense for one simple reason: Arizona was going through its hottest summer on record, and having lived there, summer life (as least from 10 AM to 9 PM) tends to consist of transitioning from one air conditioned environment to another via an air-conditioned car, with an occasional trek to a (rather warm) backyard pool. In contrast, with milder Fall weather, residents transition toward a more outdoor lifestyle. While being a battleground state didn’t help and a pulse was added for election day, the added “flu season” bump was less than for USA. But model prediction trends suggest that Arizona is still part of the surge, with one possible explanation being some reinfections.

#### Wisconsin

The Wisconsin model itself had some minor adjustments since the population is more obese that the national average (with exception for parts of Milwaukee and Madison). Summers tend to be highly celebratory, with festivals and pubs a part of the lifestyle. It is also a politically polarized state, represented in part by a republican legislature and a democratic governor. Notably, the state supreme court knocked down directives of the governor in mid-May, and while there was not a dramatic period of exponential growth, the infection curves I1 and I2 simply did not decrease enough in May-July to levels as low as many Northern states. There was also a spike in college towns in early Fall, and from May-October there were 5 Trump events. The dramatic increases of October-November where driven by rural counties, especially starting in the central-north counties around Green Bay. This gradually spread southward, county by county. The final simulation curves and metrics (**Figure 12**) suggest that with Rtot rising to 20%, curves may drop somewhat.

#### Oregon

Oregon has had lower numbers of cases than most states, consistently in the bottom 5. There have been some mild increases, often associated with targeted outbreaks in different parts of the state (e.g., veterans’ home, packing plants, church event, near Boise ID). Directives have been stronger than most states, especially for the 3 counties making up the greater Portland area, where Phase 1 restrictions remain in effect, but with some adjustments that are mostly tied to fluctuations in cases. Outdoor protests, especially in Portland, did not generate spikes, and this author’s observation at several events was overwhelmingly high use of masks (see 56). The estimate of a relatively large S and small R’s suggests risk of exponential growth, and November saw the start of such a rise, causing the governor to add CMDs. The two overplotted curves in **Figure 13** differ dramatically, based on whether the governor’s new directives are followed.

#### Synthesis

As is clear from **Figure 6**, there are considerable differences between these three states, making them good choices for comparison. For Arizona, which got hit hard during the summer, the prediction was that nearly 20% became resistant, but that decreased to ∼10% as some population shifted to being susceptible again, and now there is a second peak. Wisconsin fluctuated at moderate levels for quite a while, then got hit really hard this fall. Both Arizona and Wisconsin were “battleground” states with considerable polarity, and results were influenced by a mild added pulse weeks before the election and then a more major pulse election day (reflecting mostly the *u*_*TB*_-type voters). However, the model predicts that the Wisconsin peak may gradually subside, as the number in the R_tot_ (resistant) subpopulation is over 20% (thus susceptible under 80%) and most will remain in this status for many months. Oregon is interesting, as it has done so well. But with R_tot_ below 10% and S above 90%, it is vulnerable to a continued exponential rise phase, short-term, although strong new CMDs are now in effect for this reason. It also benefits from a slightly healthier, lower-risk population demographic.

Notice the high oscillation for the E→I1 (highest rate), and the subsequent mild daily oscillation of the I1 signal.

### 2.6 Other Example Runs: Mostly Unsuspecting Spreaders and Re-infectors, Idealized Nursing Home

**Figure 7** provides results for a run with a conceptual USA sub-population of younger and middle-aged adults, roughly under age 45. Here we just changed some model parameters to make the I1→I2, I2→I3 and I3→D fluxes lower by half (rounded down) and all the I*→R*raised by 10% (rounded up), and didn’t even change the inputs (USA defaults) that drive the model. We then check sensitivity by making a small shift of *prop*_*TB*_ (see **Equ. 2**) from 33% to 36% and overplot the 2 runs (it should be intuitively clear to the reader which is which). We can compare these results to the overall USA results while also essentially getting introduced to the empowerment provided by sensitivity analysis while using forward dynamic simulations. With R1→S not lowered, this could be viewed as the upper limit (or “worst possible”) situation for reinfections, illustrating possibilities.

**Figure 7:**
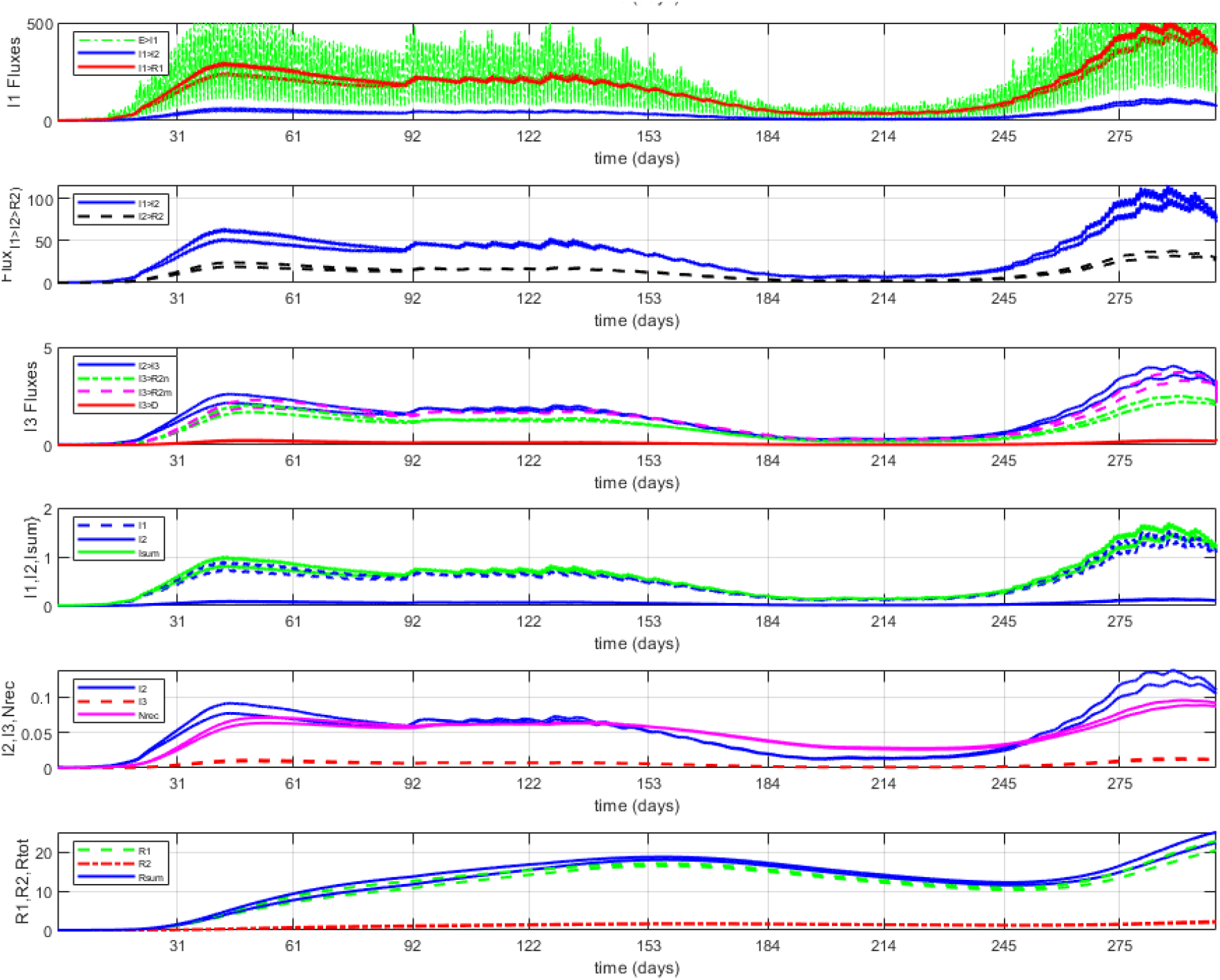
Young/Middle-aged UoD simulation. Selected plots from a “starter” model (Case 5 in default Matlab code) simulation consisting of a subpopulation composed of young and middle-aged people (see text). Notice the relatively larger I1 and R1 curves (e.g., vs I3; D doesn’t really show it, but some deaths were predicted).

As seen in the results for this rough model, more of this subpopulation does go through the I1-→R2 path, with fewer I2 and I3 complications and especially deaths. Notice that the total R population is above 10% by 2-3 months, peaks at 5 months (early August) and then starts to fall to nearly 10% as population shifts back to S, before rising again. S drops to 80% in August and then below by November (it is nearly 1-R_tot_, since total infections are mostly below 1%). The high R in summertime helps explain the lower I_tot_ levels in September-October. If the proportion of S that has gone through the infection-recovery cycle is assumed to have the same likelihood as other S for exposure E (i.e., well-mixed), with this assumed model not only is reinfection cycling common (estimated at about 3%), but that by the end of the 9 months, 44% of the population has been infected and over 25% has cycled back to the S population. These are likely high, as the flux adjustments in the model were probably too extreme, but nonetheless, it illustrates a possible challenge. Also, from the overplots it is clear that having a greater proportion exhibiting *u*_*TB*_ behavior has a detrimental effect on the “common good” of this population, and of course, those who are more vulnerable to which they may actually come into contact.

**Figure 8** provides an example of an “enclosed/isolated” population, with the UoD here a large hypothetical nursing home idealized to a population of 1000 persons (mostly residents). As it is an older population with health risks, some changes are made to the model fluxes, mostly decreasing I1→R1 and I2→R1, and increasing I2→I3 and I3→D. Additionally, the relative “frequency” coefficients within the mapping matrix (**Equ. 7**) were made higher (e.g., I2’s still in contact). The simulation started with the assumption that at *t=0* a caregiving worker (i.e., 0.1% of population) unknowingly has an asymptomatic infection I1 that is not discovered for 3 days (during which they provided caregiving and interacted with co-workers), enabling a degree of spread. Soon thereafter we assume a strong CMD lockdown, maintained for the rest of the 100-day simulation. As seen in **Figure 8**, I2 (symptomatic) peaked within a month, and most deaths were between about 2-8 weeks.

**Figure 8:**
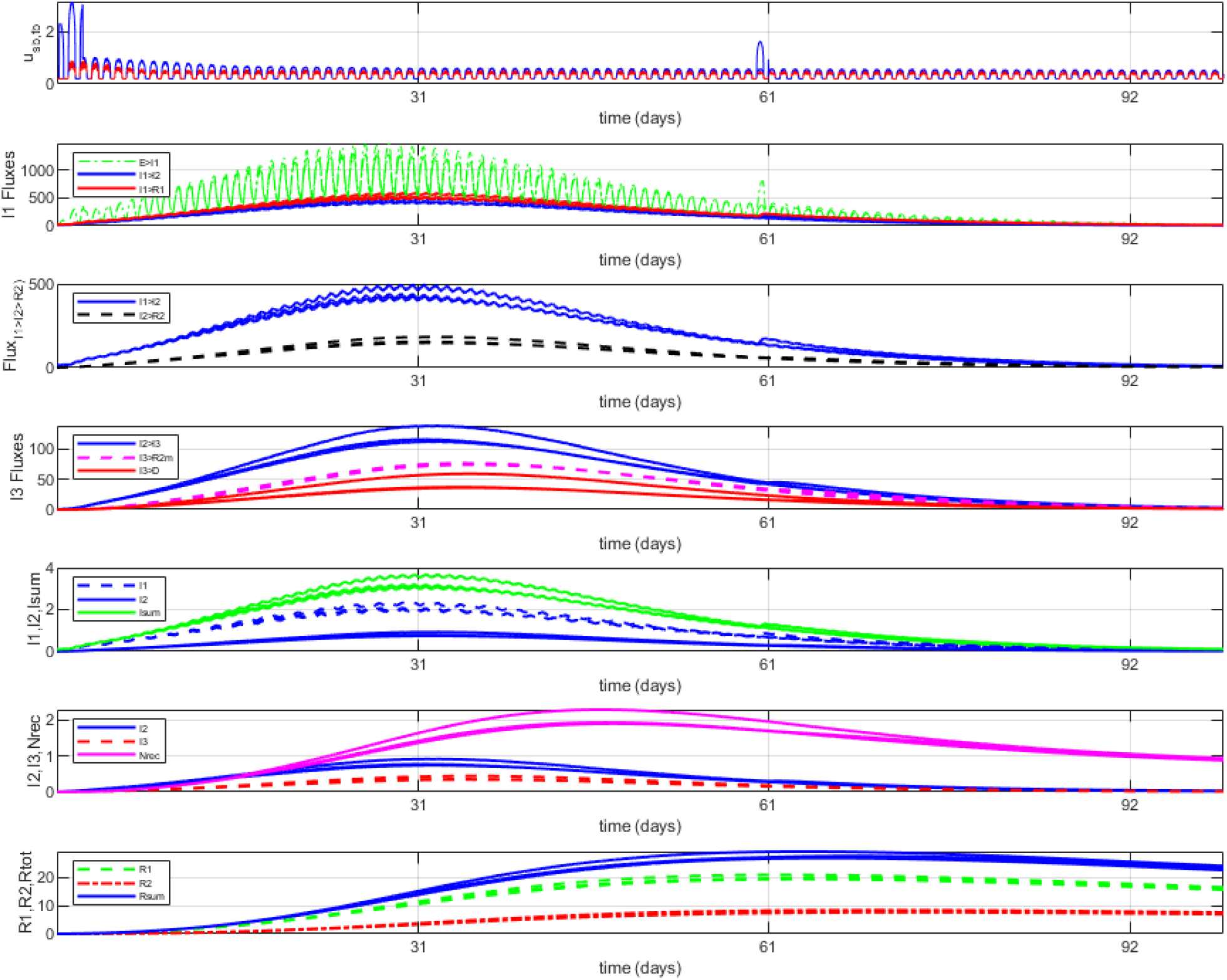
Idealized UoD for Nursing Home Simulation. Selected plots from a “starter” model (Case 6 in default Matlab code) consisting of a subpopulation composed of the community within a nursing home facility, here idealized to 1000 persons (mostly residents), for a 100-day simulation. We assume a caregiver (1 of 1000) works 3 days as I1 (asymptomatic), interacting with residents and co-workers. Then within 1 week aggressive measures are taken, and maintained (to letting *u*_*SB*_ = 0.5). Two cases are overplotted that differ only by medical intervention: the higher curves are for an “April”-type situation where medical COVID-19 knowledge was still evolving (*u*_*i1*_*=u*_*i2*_ = 1.1 to 1.2), then an “October”-type with hospitals below capacity (*u*_*i1*_*=u*_*i2*_ = 1.4 to 1.5). In the first, 25 people (2.5%) are predicted to die, while in the second, 16 (1.6%). Also, overplotted for the second case is a new 1-day I1 “situation day 60” that causes only moderate effect (as strong CMD’s are in effect).

Overplotted are 2 scenarios for medical interventions (e.g., April with less COVID-19 experience and a full hospital versus October with more knowledge and space); the predicted deaths dropped from 25 to 16. Notice that: the number of infected reached a peak of 3-4% at about 1 month, about 1 in 25 were predicted to be hospitalized, and over 20% were in R (resistant) within about 40 days (includes workers).

### 2.7 Longer Trend Predictive 400-Day USA Runs, With Vaccines, Differing Immunity Assumptions

Vaccines are arriving that appear safe and effective, but at this stage with unknown time windows of immunity. It is useful to use the model to predict possible consequences, given certain assumptions, of combinations of vaccine administration and CMDs. Here we continue the 300-day USA simulation to 400 days, augmenting these two scenarios with several assumptions on vaccine longer-term effectiveness (e.g., with the shorter S→R1→S path being 90%, or the longer S→R2→S path being 90%). The latter also represents the possibility of longer immunity, given there would only be up to 100 post-vaccination days in this simulation. Vaccines are assumed to be administered gradually to the population, with a lower initial dosing rate gradually starting in mid-December followed by the second “booster” 3-4 weeks later adding effectiveness, with 2% of the population inoculated by the end of January, 10% by end of February (365 days), and 20% by Day 400 (early April). During this time there continue to be two alternative CMDs adherence alternatives, but with the “flu season” bounce diminishing.

The results illustrate that CMDs still matter (see also **Figure 5**, but here with “Christmas pulse” added, especially for the less-compliant alternative. Notice a peak in early January, so high it compresses shapes of other curves. With this worse scenario, I_sum_ infections reach nearly 2% as the new year starts, but still pass 1% for the other, and for both R_sum_ crosses 20% by January. The two alternatives are very different until March, but less so as the “flu season” bounce diminishes starting in January (complete by April). The vaccine augments these predictions, but ironically it is subtle on these high scales. With vaccination, we end with over 30% of the population within *R*_*sum*_ and rising, and S is below 70%. A key prediction is that infections will drop by springtime in any case, but more so with vaccinations (hard to see on this scale). These results strongly support the priority for administering vaccines first to vulnerable populations and key at-risk essential service providers such as healthcare workers, as the predicted next “peak” wouldn’t be until summertime even without a vaccine.

## 3 Discussion

One overriding finding just documents the obvious: I1 (asymptomatic or very mild) not only matters for this pandemic, but dominates the rather sensitive model results, both for SARS-CoV-2/COVID-19 decay and rapid growth. The primary driver is S↔I1 encounters, with I1↔I1 and S↔I2 next but comprising well under tenfold an impact (except for certain idealized UoD’s such as nursing homes). This suggests the need for massive, **ubiquitous surveillance testing** to find and track asymptomatic individuals. Measured “cases” still appear to represent more I1→I2 fluxes (i.e., showing symptoms) than S→I1 (mostly asymptomatic). Model results for the USA are consistent with recent synthesis that roughly 10% of the USA population have antibodies (e.g., CDC); the model predicts this will grow slowly, as members of the R_sum_ subpopulation transition back to S (susceptible).

The results do not seem consistent with the CDC’s continuing advisory that reinfection with COVID-19 is rare. At issue is the “mild hidden” I1→R1→S path. Perhaps the model outflux parameters from R1, informed by the emerging (but somewhat conflicting) body of knowledge that antibody protection especially in mild cases drops significantly within 2-6 months (48-52) and even more so in older persons (51), have flux values that are too high (average is 0.009/day for R1→S). But this seems consistent with physiological expectations, as antibodies are a class of proteins that should mostly break down within a few months. A longer half-life implies targeted adaptive memory B and T cells that are tuned and upregulated. There is some recent evidence that this may be happening, especially for B cells (52-54). The consequence is considerable. If the R1 and R2 outflux parameters are dramatically lowered, the closed-loop SEIRS-type model essentially becomes a feedforward SEIR-type model where R is a population collection sink (a dream scenario). In such a case, the susceptible (S) population would be rapidly dropping below 80%, the R subpopulations would be rapidly rising above 20%, and *u*_*TB*_-type people who have had the virus can feel free to “go wild” since the predictions of **Figures 6-7** are less of a worry.

A similar concern emerges with the upcoming vaccines, especially for timed 2-bolus mRNA-based and protein-based vaccines, as to how long the associated immunity will last. The model currently represents a generic vaccine as a flux that can be distributed between R1 and R2 (**Figure 9**), which starts to matter even within 100 days. The model also has one well-mixed S subpopulation, assuming no large-scale trend toward reinfection(s) being either more mild or worse than the original. It would straightforward to add a second S state.

**Figure 9:**
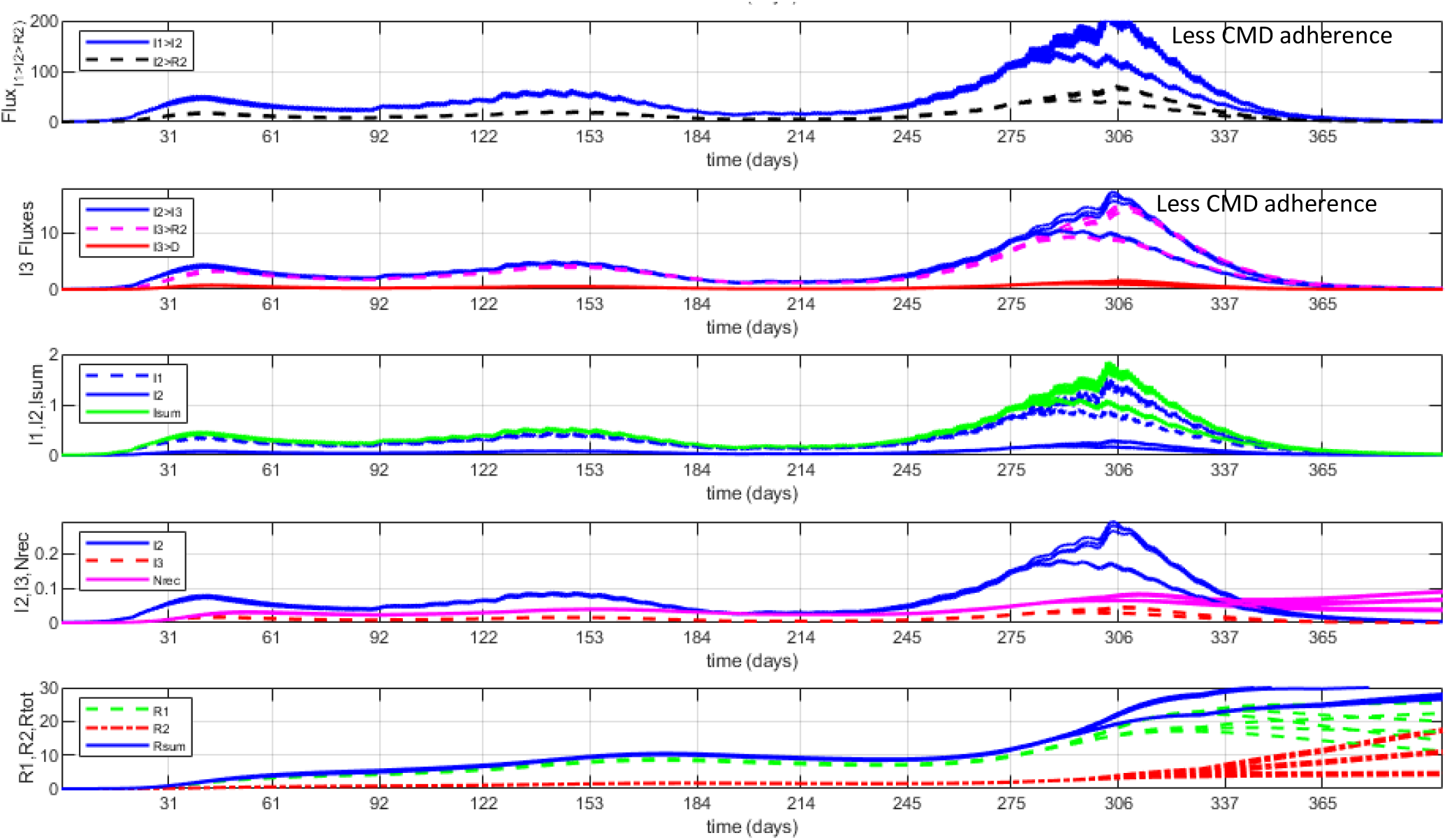
Extension of the Figure 5 (USA) simulation to 400 days, including effects of start of vaccines. Selected plots from a “starter” model (Case 11 in default Matlab code), with the 2 scenarios of **Figure 5** continued, only with the added “flu season” signal diminished by April and administration of vaccine ramping up, here assumed administered to 10% of the population by March and 20% by early April. Overplots include vaccine weights for S→R1 vs S→R2 flipping 90%-10% each way, best seen via the (rising) R2 signal (dashed red). With vaccines, R_sum_ crosses 30% (and rising), and S ends well under 70%. Infections stay very high through all of January (compare to the “now small” previous two surges), and new deaths peak in early January.

What follows assumes that there will remain challenges for most of 2021 and perhaps beyond if the pandemic transitions into an endemic (50). It also assumes that there will be a significant subpopulation that falls into the “not recovered” category, with various new acute and chronic health challenges that deserve study.

### 3.1 Added Value of Dynamic “Phenomenological” Models with Embedded Signals and Fluxes?

An added value of dynamic models with **internal structure**, versus based on curve fitting, is that internal estimates of signals and fluxes are available. These include some that relate to measured data, but importantly, some that do not. In control engineering, models are routinely used for “state estimation” – using measurable outputs (and sometimes inputs) to estimate “under the hood” internal phenomena (e.g., “states” and state-dependent ancillary signals such as fluxes) that enable better control of a system. This is a common value-added of dynamic models and has been an important tool in control (and biomedical) engineering since the 1960s.

As an example, we have data on both hospital admissions (i.e., here ∼I2→I3 flux) and hospitalizations (i.e., here associated with most of I3, and a small subset of R2 that are still recovering). If these track well in the model, the chances are pretty good that I1 and I2 will also track well, especially since consequent estimates of R1+R2 (“resistant” subpopulation) also seem consistent with CDC and recent estimates (e.g., 51,55).

It should be noted that while the structure of the model has been reasonably refined, the current model-tuning by this author is just a start. But with the model displaying **reasonable temporal dynamics** and being so **sensitive to flux parameter ranges**, it does not seem likely that thorough refinement will produce major changes in parameters. Hopefully teams with more resources will give refinement a try. Even so, some insights that seem to be of value have emerged. Specifically, since model results are reasonably synergistic with measured metrics, this suggests that **internal signal estimates** may have value. For instance, for the 10-month USA and 3 state simulations, runs using the default model suggested insights given in **Table 2**.

Perhaps this type of dynamic model will be especially useful during 2021, as **vaccines** gradually emerge (here supported by slope inputs) that are likely effective (e.g., perhaps 95%), at least for lowering the risk of COVID-19 disease. But whether these prevent small infections in and transmissions by inoculated persons remains an issue. Another issue is whether vaccines will provide only seasonal immunity for this type of coronavirus, in which case the current **pandemic** may gradually evolve during 2021 into an **endemic** (50). If so, tools will be needed to help **optimize this balancing act**, especially given a polarized society. To the engineer, this is a classic optimization problem where an overall “performance criterion” must strike a weighted balance between the **benefits** of diminishing the COVID-19 disease and the associated personal and economic **costs** to society.

### 3.2 This Model as a Dynamic Tool Assisting Studies “Above the Hood” and Forms for Higher-Level Models

The SEIRS-Nrec model needs to be driven, here by use of the concepts of encounters and medical interventions. Actual drives are clearly the fruits of manifold agent-based CAS processes, both within society and within our bodies. Valuable studies of social behavior as related to COVID-19 range from mobility (23-30) to impacts of mask use (2,32-38). All have merit, and such research was are not intended to help drive an epidemiologically- and medically-oriented model. But perhaps interpretations of such results could be broadened and/or refined by driving a dynamic model that also estimates internal phenomena, including in near-real time. The SEI_3_R_2_S-Nrec model might serve as a tool to assist **ongoing interpretation** rather than as the primary research focus.

Research can help inform social CMDs such as the form, timing and priorities of lockdowns. A modeling challenge, though, is to distill such information into a form that can drive a model, which here implies mapping to the concept of **interpersonal and biophysical** encounters. In reality “encounters” is more of a biophysical than a social construct. For example, a person who enters a crowded bar with marginal ventilation has a potential “encounter” with every person in that bar. The “encounter” between a masked person and a masked employee while paying for groceries is quite minimal, as is the encounter of masked persons in an outdoor setting at a Black Lives Matter protest (56). This concern provided motivation for having *V*_*high*_>V_low_ for E→I1 but not for the other outflux rate of E→S. In this way, as the E signal became higher, the likelihood of E→I1 grew. But **compartment E** is ripe for becoming a biophysically-oriented submodel with internal dynamics (**Section 4.4**).

As an example, consider this author’s iterative thought process in coming up with slope and pulse sequences for runs. It was heuristic, as suggested by **Figure 2**. Essentially this author was trying to become an “expert” in a new field, but with the benefit of decades of experience with other types of bio-models and biomedical projects. “Data” to be heuristically integrated fell into two types of information, extracted over time windows that could reach 9 months: COVID-19 datasets on the one hand and a diversity of ever-changing social directives and degrees of societal adherence on the other. But we’ve noted that available data that was not a perfect match to certain signals and fluxes. Further, there was the challenge that both **response capacity and knowledge evolve** over time. Consider these examples of the thought process (e.g., **Table 3**) used by this author for simplifying incomplete data and CAS behavior into tractable (yet heuristic) drive signals (here mostly based on USA design).

**Table 3:** Examples of how synthesis of heuristic concepts can lead to (fuzzy) if-then rules.

|  |  |
| --- | --- |
| *Thought prioritization on **early months**:*<br>Because of lack of adequate testing capacity and even hospital capacity in some regions, "cases" were dramatically lower than what was actually within the population, including for both asymptomatic and even symptomatic, for the early months (e.g., March-April-May-June). Further, while hospitalizations and deaths were better metrics early on, in some heavy-impact regions people became seriously ill and sometimes died outside of hospitals. Outbreaks were more regional, and directives diverse. Further, early "lockdown" directives, while aggressive, were suboptimal, as organizations such as WHO and CDC hadn't yet acknowledged the scope of airborne aerosol transmission and essential need for universal mask use (1,2,31-35). | If during early months:<br>{I<sub>3</sub> is just over reported hospitalizations} and<br>{D is close to reported deaths} and<br>{I<sub>2</sub> is higher than "cases" data},<br>Then {model results are adequate}<br>If {initial directives high in group sacrifice} and<br>{early effectiveness is only moderate} and<br>{degree of "pandemic fatigue" by June},<br>Then {$u_{SB}$ & $u_{TB}$ ramps drop no more than 40%}<br>and {$u_{TB}$ pulse events start Memorial Day}" |
| *Thought prioritization **summer months**, for ramp and pulse events:*<br>Gradual reopening for the summer was all over the map, as was adherence. There was dysfunctional national leadership. Spikes had many reasons, from bars/restaurants to nursing homes to packing facilities to worship gatherings to the Sturgis event, and tended to be regional. One commonality is 3 major holiday weekends (starting with Memorial), and weekends in general – these could be pulses, and often these did not follow directives. People in very warm climates (e.g., Southeast, Southwest) are more likely to spend time indoors during the summer, adding exposure risk. Many states added directives in late Summer to manage spikes. Opening/closing of schools and businesses was mixed. Mask use scientifically recognized as priority, but polarizing. SARS-CoV-2 testing, while still suboptimal, had grown (with some asymptomatic cases), as had contact tracing. Medical interventions gradually more effective (e.g., reducing deaths). | If {spreader pulse events have no clear pattern}<br>and<br>{a modeling goal is to illustrate cause-effect},<br>Then {add USA pulses for holidays and weekends for $u_{TB}$}<br>If {reported cases is slightly lower than $I_1 \rightarrow I_2$},<br>and {D is close to reported deaths},<br>Then {run is more-or-less satisfactory}<br>If {fewer relative deaths}<br>Then {$u_{M1}$ slope can increase} |
| *Thought prioritization for "**flu**" and **election** season:*<br>Colder weather both brings people indoors, and naturally increases risks of colds and will certainly affect respiratory transmission. Fall first hits in Northern states (starting with Midwest, Mountain). Special events included in gatherings in election battleground states, and voting. Polarized nation, and election implies more rallies and voting lines. As cases and hospitalizations surge, added directives return for states, especially mid-November. | If {weather is cold} and {more are inside},<br>Then {add seasonal effect to $u_{SB}$ & $u_{TB}$ slopes}<br>If {country is deeply polarized},<br>Then: {divergence between $u_{SB}$ & $u_{TB}$ is more} |

### 3.3 Could Advanced Tools Like Fuzzy Expert Systems Distil Guidance and Help Inform Consensus Directives?

One strategy for integrating such a heuristic mix of data and expert knowledge that is hard to quantify into a formula is to develop a Fuzzy Expert System (FES), an AI tool that has been used since the 1970s (including for medical diagnostics). The heuristic “rules” given above in **Table 3** are examples, where the phrases within brackets include a fuzzy signal associated with a membership function. The idea is to synthesized knowledge into a collection of “soft” rules that have degrees of “truth” from 0 to 1, with coding protocols for how to integrate “consequents” for different types of rules. While during this past decade the AI approach of machine learning has especially blossomed, the two are often combined (4,30), for instance in this author’s co-development of a “neurofuzzy” package called SoftBioME (Soft Biomedical Modeling Environment) for use as a research and educational tool (57). Bullet points such as in **Table 3** could potentially be mapped into fuzzy rules.

In fields of medicine, **clinical practice guidelines** are common that are intended to provide consensus on best practices. These are based on synthesis of research and clinical expertise, usually by a panel of experts. Whether formalized or not, such guidance is essentially part of the aim of organizations such as WHO and CDC, and as we have seen, it tends to be cautious, but will nonetheless evolve. All guidance tries to simplify CAS behavior into sometime more tractable, such as using covariates (11) with mask use and limiting indoor social gatherings as a top two, then hand/surface cleaning, social distancing, context-specific quarantine times, etc.

A FES is just one tool for coding (and updating) such knowledge, and its output could be targeted differently to different stakeholder groups. An advantage is that the FES structure tends to be robust to the inclusion of added rules (4,57). Results, while numerical, can be mapped to weighted suggestions for robustly responding to a given context as it emerges. One advantage might be to help as a dynamic resource for local officials needing to fine-tune local directives that are responsive to ongoing changes, such as a targeted new spike in cases.

### 3.4 Roles for Control Engineers in Helping Optimally Manage in Real-Time the 2021 Pandemic Response?

For well over a century, modern technological progress has tended to follow this progression:

basic science → applied science → engineering → technician support

Scientific “handoff” to engineering is common, often with continuing iterative R&D efforts as products and services evolve. Over this past year scientific knowledge of SARS-CoV-2 and COVID-19, building on an existing knowledge base, has evolved considerably. Furthermore, novel types of expertise, including from the “big data” community, have sought to help the public health community in addressing this global pandemic. But the above progression has not been followed. Perhaps we’ve replaced “engineering” with “politicians” and technicians with contact-tracers. While effective political leadership can make a difference (e.g., in New Zealand), the **problem-solving skills** and approaches of engineers may add value. This author suggests that engineers and other technical talent may have a strategic, and largely untapped, role to play in helping the public health community in effectively managing the pandemic into 2021, acting in near real time.

**Figure 10** presents a block diagram structure, likely recognizable to any engineer who has taken a “controls” course, that consists of a real-world dynamic system to be controlled, a model in parallel, an “intelligent” feedback block, a controller, and a feedforward block. It is intended to **operate in real-time**, with feedback of new output information helping a control algorithm adjust a control strategy to enable robust performance. The idea is to make informed decisions that include processing of new “output” information, perhaps augmented by “state estimators” (of “under the hood” phenomena) if using a model such as SEI_3_R_2_S-Nrec. Engineers with augmented educational (or on-the-job) experience in “optimal” and “adaptive” control are well-trained in strategies that balance competing objectives and in how to adjust responses as situations change.

**Figure 10:**
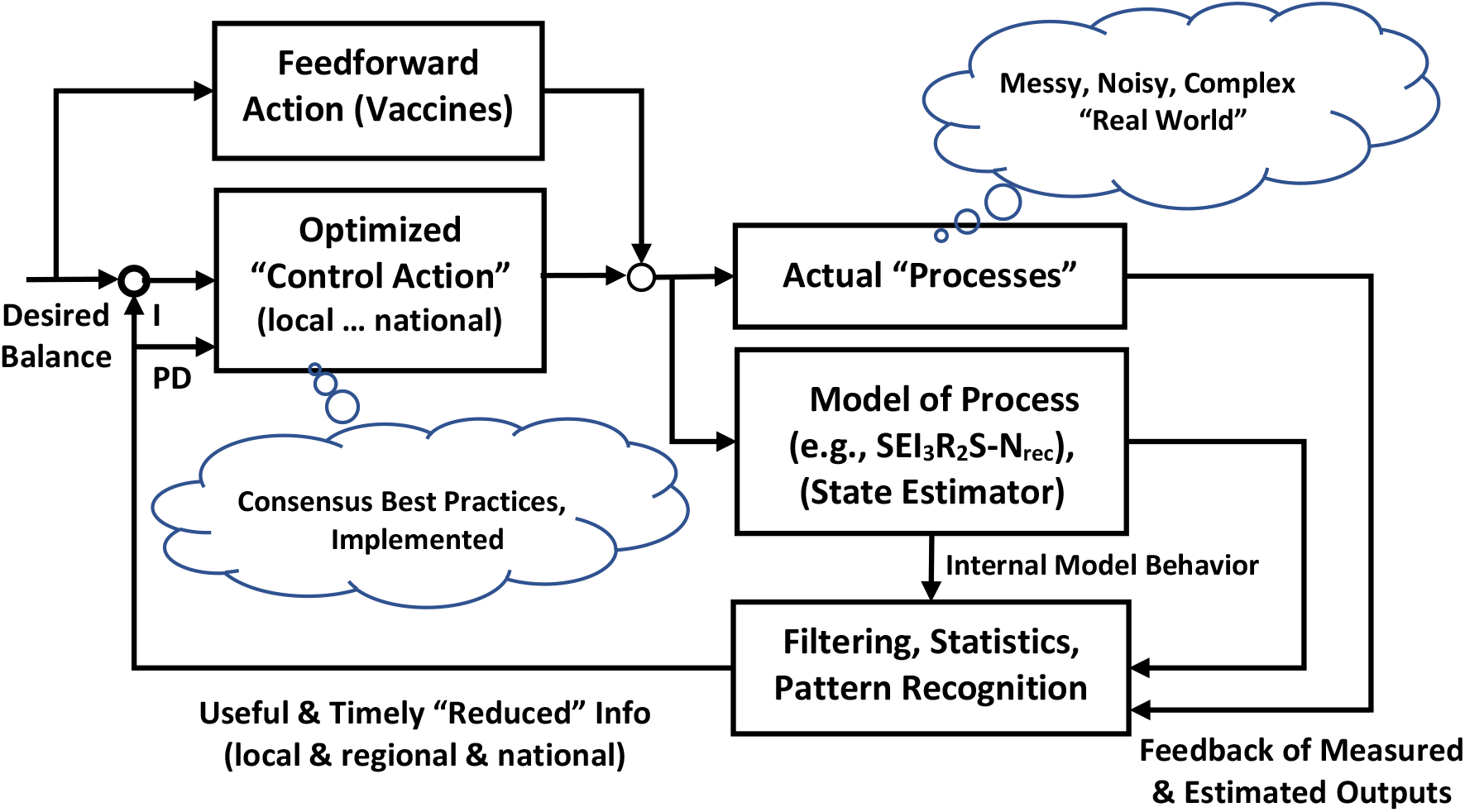
COVID-19 real-time management strategy from an idealized “systems” perspective. Here a model such as SEI_3_R_2_-Nrec is set up to act in parallel with the “real world” model as a state observer for estimating unmeasurable phenomena, which in turn can help with “intelligent” filtering and pattern recognition, whether by traditional or newer AI approaches (which also can be used to adaptively tune the model, not shown). Reduced data (and updated risks, 29,43) is fed back for use by control action. Here PD represents” proportional-derivative” type control action, where P implements a proportional response (e.g., responsive temporary adjustments in CMDs) while weighing competing subcriteria, and D action represents rapid transient response to changes (e.g., dependent on comprehensive testing rates, timely contact tracing and anticipatory testing, with I1 focus), usually fast-acting but moderate in scope (“gain”). Here I-action represents a “leaky integrator” (integrates away performance error but not at all costs) that aims at balanced strategies over a longer time horizon (e.g., lowering metrics like cases, hospitalizations, and death while also addressing societal costs, where here the “future” is months). Vaccine delivery is viewed as a feedforward strategy, with limits on speed of delivery and prioritization of resources. If feedforward action is limited (e.g., temporary immunity), a well-tuned feedback path (e.g., testing-based, timely directives, adaptive) still can provide robust performance. Such a conceptualization may be helpful for 2021, perhaps longer if the pandemic evolves into an endemic.

In this conceptualization, imagine engineers reporting to a local public health expert, and managing a team that includes contact tracers and data-driven testers. One side advantage of engineer involvement is that unlike scientists and politicians, engineers tend not be “polarizing” to the general public. For instance, if armed with data and modeling predictions, CMDs could be more precise and targeted, such as targeted closings for the minimum possible days (conditional on new data), with transparent decision-making based on ongoing data.

### 3.5 Public Health and COVID-19 Disease: Reasons for Expanding Nrec (Not Recovered)

Public Health is a field and profession that encompasses much more than epidemiology. It also includes health indicators of functional impairments and dysfunction, health disparities, and other indicators of quality of life. We’ve noted that the disease of COVID-19 is creating a portfolio of health challenges, especially damage to tissues. Whether due directly to SARS-CoV-2 activity or to consequences of an aggressive immune/inflammatory response, the extent of associated functional impairments and dysfunction remains in area of ongoing research. Affected tissues include respiratory/pulmonary, cardiac, renal, intestinal, neurosensory, and neurocognitive. These in turn can cause measurable organ dysfunction. Some appear to be acute and usually recover (e.g., smell, taste), while others have the potential for being a sustained chronic condition (e.g., respiratory).

This author, with a heritage more in physiology and systems biology (mostly as related to rehab science and engineering) rather that epidemiology, found it somewhat painful to reduce the immune system – one of the top bio-examples (along with the brain) on virtually any complexity scientists top list of bio-CAS examples – to a few rate constants. But while this made sense here, there are natural public health ties to degrees of recovery, for fields ranging from respiratory therapy to rehabilitation medicine to disability services.

The Nrec placeholder model could be expanded and partitioned to addresses types of tissue damage in the context of the immune response, then the repair and remodeling process and roles for interventions.

### 3.6 Possible Future Modeling Directions

The initial plan of this author involved developing a collection of SE_3_R_2_S-Nrec “personas” that would operate in parallel, intended to provide representative diversity within populations that went beyond the usual unimodal distributions that (to this author) made less sense for COVID-19. Personas would interact only via “encounter” interfaces. It was driven by concerns of **diversity** across both behavioral and physiological domains: ***i)*** well-documented of risks in older adults (e.g., 39,40) and yet profoundly less in children (e.g., 41, but see 42) who may even possess helpful antibodies (e.g., 54), ***ii)*** diverse preexisting conditions ranging from obesity/diabetes to racial disparities (e.g., 29,45), and ***iii)*** clear recognition of what constitutes super-spreader behavior. Further, coding into “persona” arrays (29,31) is straightforward (although estimating the many parameters less so).

This is now viewed as not the best idea, at least here. The reason is that a better approach for implementing (non-unimodal) heterogeneity and diversity seems to be to provide context-appropriate diversity and/or statistical variance **within key compartments**. This follows the approach broadly used in biochemistry, where “biochemical species” react within a conceptual or physical compartment (e.g., “bioreactor”); well-tested software packages such as COPASI (58) illustrate this approach. Compartments can have **internal dynamics**, represented by more than one state. For example, compartment E could contain a biophysical submodel with faster internal dynamics that aims to study cell invasion and viral load translocation dynamics using the respiratory and circulatory flow systems, and I3 and R2 could have different sub-populations partitioned by health risk factors. The cleanest approach is for fluxes between compartments to serve as “dynamic constraints” for the internal dynamic model. If there is a desire to pass “risk personas” between compartments, there can be subfluxes within a connectivity matrix (12,17,18), but this can lead to parameter explosion. This is best pursued by multidisciplinary teams having the targeted expertise needed to create and evaluate such submodels.

Because the current model is inherently **asymptotically stable** (a consequence of being a “closed” system with inputs only modulate fluxes rather than deliver power (or people) to the system), it is best if sub-compartments are “closed” as well. This differs from classic predator-prey (and some host-parasite) models which are often designed with a “carrying capacity” based on limited resources (e.g., hosts-as-resource) (6), or physiological models containing regulated “capacity parameters” based mostly on evolved (and adaptive) size-scaling (57).

Still another plan by this author conceptualized an AI-inspired FES approach (4,30), building on past work with SoftBioME (**Section 3.4**), which can produce nonlinear conserved fluxes and ODEs based on causal “changes” intuition. But as suggested by **Figure 10**, FES packages are now seen as supportive of **control action** designed to minimize high-risk encounters via the *u(t)* drive to model, just as **AI machine learning** (e.g., pattern recognition) approaches are best seen as helping manage and distill incoming information via the **feedback path** in **Figure 10**.

Perhaps the most intriguing future direction, at least to this author whose background is more in physiology and rehabilitation, is to **expand the Nrec submodel** to try to capture the types of pathophysiology and functional impairment known to be associated with the COVID-19 infection and recovery bioprocesses. This includes degrees of functional impairment (e.g., of tissue types), possibilities and limitations for tissue repair and remodeling (both spontaneous and with medical interventions), and degree of dysfunction (e.g., of organs). Such models could be weighted mappings based on statistical data (thus new outputs), but more likely new dynamic states would be needed to help better understand the transient nature of acute and possibly chronic health conditions. FES provides one approach for expanding the Nrec submodel, especially by integrating the diverse range of evidence into rules that partition outputs and states into types of tissue and organ dysfunction, and then relate these to degrees of impairment and disability. In such models, “acute” phenomena are states, while “chronic” conditions are either output accumulations or “slow-leaky” states with very low outflow rates.

Finally, while this model was developed specifically for the COVID-19 pandemic, it could conceivably serve, with a similar structure but with different parameter tuning, as a robust tool for **other classes of infectious diseases**, especially those that include both epidemiological and medical intervention challenges.

Going full circle, it could also serve as an **educational tool** for helping students to better grasp bio-phenomena through use of causal nonlinear bio-models, as was the original intent before this project grew and evolved.

## 4 Methods

### 4.1 Model Structure: SEI_3_R_2_S Epidemiological and Nrec Medical Submodels, Coupled

The core epidemiological model structure network, summarized in **Figure 1**, extends the classic serial chain SEIR model by introducing a lattice of connectivity, resulting in the SEI_3_R_2_S epidemiological model. Here the I1-I2-I3 compartmental states represent degrees of infectivity. Through the lattice design, a subset of population transitions directly to R1 (shorter, mild “resistant” phase), thus bypassing infection levels I2 (i.e., clear symptoms) or I3 (i.e., ideally hospitalized). These transitions, represented by arrows, are fluxes with units of population flow transition per time (here in “per day”).

#### Epidemiologically-motivated SEI_3_R_2_S submodel

As in traditional for SEIR models, “flow” is only unidirectional. In engineering science bidirectional models are also common where downstream can affect upstream (e.g., electrical transmission lines, pipes with fluid flow such as the circulatory biosystem, rivers without waterfalls). Then flow is from the higher state value to the lower value, which can be more physically intuitive. In contrast, here “downstream” compartments (e.g., R’s) can hold more population (see **Section 4.3**).

A key systemic property of such idealized fluxes is that the amount of **population is conserved**, i.e., the outflux from the upstream equals the influx to the downstream and thus subpopulations are simply redistributed over time (see ***Equations 3***). Thus compartmental states 1-7 sum to a total of 1.0 for each timestep, except for any deaths that accumulate through an outflux to the environment. For better visualization of the ODEs, when possible an outflux in one ODE equation is stacked vertically with the associated influx of another. All signals *x*_*i*_*(t)* and *u*_*i*_*(t)* are a function of time (though for easier reading this is not shown explicitly here):

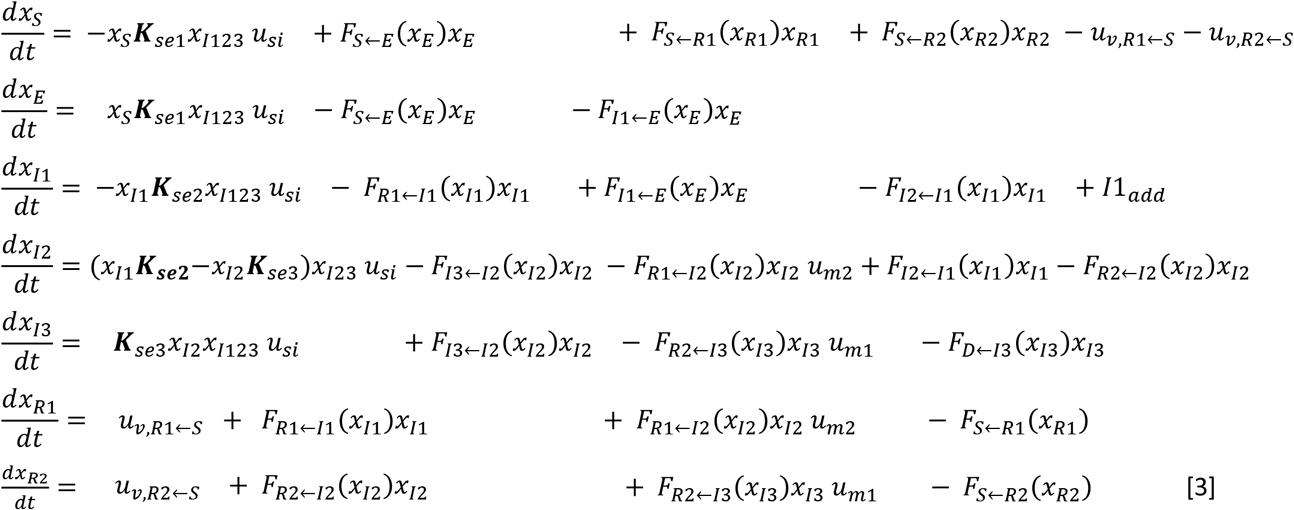

where *F*_i<j_*(x*_j_ ) are Hill rate functionals (59,60) from state *I* to *j* fluxes, *K* is a weighing vector for input-modulated 2-state encounter effectiveness (see **Equ. 7**), *u*_*si*_ is the overall “encounter” input, *u*_*m1*_ and u_*m2*_ are medical inputs that multiplicatively modulate key rates (i.e., improving outcomes), *u*_*v,R*_*←*_*s*_ vaccine fluxes, and *I1*_*add*_ importations.

The concept of an encounter “strength” matrix (***K*** rows) seems to be a novel concept, but reflects the reality that with three I’s, S can be exposed to I2 and I3 individuals (e.g., clinician in hospital). Also, an I1 individual can not only interact with S but also be exposed to other I1’s (e.g., in social setting such as a bar) or other I2 or I3 (e.g., a loved one), thus increasing their chance of transitioning to I2. Each coefficient is contextual, a multiplication of “effect” and “frequency” (**Equation 7**). In simulations, S↔I1 encounters dominate for larger populations; but other encounter possibilities can influence smaller UoDs such as a hospital or nursing home.

#### Medically-motivated “Not Recovered” submodel

The 8^th^ state, Not Recovered (Nrec), is intended as a placeholder for connecting the epidemiological model to model based more on pathophysiology and medical intervention. It is motivated by the observation that “resistant” and medical “recovery” are different constructs, with “recovery” differing both dynamically and functionally from the dynamics of the R1 and R2 states. For instance, it is widely documented that many people die of consequences of a host-pathogen “battle” within, even when the level of antibodies suggest an individual is in theory resistant (e.g., R2). Examples are extensive tissue damage (e.g., lungs, heart) due to “cytokine storms” (46) or local consequences of ACE2-related dysfunction (61). Further, accumulating evidence reveals residual impairment and dysfunction in survivors who would classify as “resistant” and then eventually as “susceptible” for reinfection while already compromised. Here the state receives strategic fluxes from the core model as driving signals and medical intervention inputs:

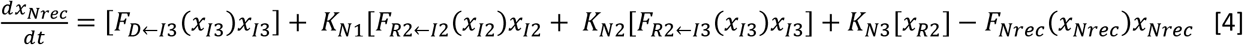

where here we assume that non-recovery is driven by **100% of deaths**, plus the **degree of not recovered** (weights *K*_*Ni*_) in survivors. The submodels should be coupled, in principle bidirectionally, but here we start with a simple unidirectional approach where the *Nrec* signal is a function of the fluxes of I3→D, I2→R2 and I3→ R2 (and thus I2 and I3), plus a small fraction of R2. Finally, the variable rate constant for decay of the *Nrec* state is, by default, smaller for low levels. This helps reflect the medical observation that many people appear to have a degree of near-chronic functional impairment and/or dysfunction, presumably owing to a degree of residual (plastic) tissue damage. Note that at this initial stage, the medically-oriented “Not-recovered” model does not break down impairment into categories (e.g., heart, lungs), but this would constitute a natural extension.

### 4.2 Types of Fluxes

In ***Equations 3 & 4*** there are two types of fluxes, both of which build on a large history of similar approaches and involve nonlinear operations:

- ***“Encounters” between 2 signals, with multiplicative input-modulated “strength”*** (e.g., see **Figure 2**)
- ***Conventional unidirectional fluxes where a rate is a function of the upstream state, here implemented using the classic Hill function*** (59,60)

As a context, for many SIR or SEIR, many of the insights are based on using constant rates, and thus assume that fluxes between compartments are linear.

The usual exception the first “entry” rate, often a multiplicative modulator called β (11) or λ (**Section 1.1**); the latter is often based on a statistical or conceptual fit (e.g., normal (Gaussian), Poisson). More advanced models use two λ’s (15), one for variance in social behavior and the other variance in some population exposure-susceptibility metric. Both modulators can operate as a simulation launch, but our approach builds on β.

Such idealized, mostly linearized models tend to aim at addressing concepts like reproduction ratio and herd immunity. Here we follow use classic “systems and controls” and “physiological modeling” approaches where the modulating input is a dynamic signal that drives the system, as introduced in **Section 1.1** and **Figure 2**.

#### Nonlinear fluxes via input modulation

This approach bears similarity to how many biosystems are modeled where biochemical species interact (e.g., enzyme reactions, hormonal modulation), as well as countless models in population biology were species interact (e.g., predator and prey). In such formulations the flux is the product of two signals modulated by another signal, of the form:

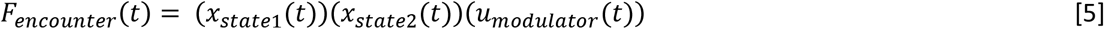

where input *u* modulates the “strength” of the encounter, and the *x*’s are the states “participating” in the encounter. Here states are on the interval <0,1>, with the S signal starting near 1.0 and I1 and I2 signals starting small but non-zero. For the infection to grow (i.e., I1 to grow), *u(t)* must start and stay high enough to facilitate new encounters. “Spikes” in the *u(t)* trajectory will cause large increases in infections (E→I1) only if I1 is already high enough for there to be increased S↔I1exposures. Since S changes slowly relative to 1.0, If we assume S is constant and *u(t)* is sustained (approximately constant) over a time window, this flux is linearized. In such a situation the model will naturally predict initial exponential-like growth in I1, although this will slow as it gradually “discharges” to signals I2 and R1.

#### Nonlinear flux rates via Hill functions

In fields ranging from physiology to pharmaceutical biochemistry to clinical medicine to population biology, “exponential decay” (e.g., “discharge” rates in response to an “initial state” or “input bolus”) and “exponential growth” (e.g., to a steady input drive) tend to be crude approximations that are applicable only over neighborhoods of time and amount. Various “systems identification” tools that check for linearity and model order find **pervasive nonlinearity**, especially varying rates. For instance, bio-rates of decay are usually higher before the “half-life” time, then much slower. Here we assume a state-dependent range of rates, from *V*_*low*_ to *V*_*high*_. There are many math operations for “soft saturating” the transition between these two extremes, including the logistic, error (erf()) and arctangent functions common in AI. But we will use a classic that has over a century of history in the physiology and pharmaceutical fields with, intuitive describing parameters: Hill functions (59,60). In deference to SEIR models where most rates are assumed constant, we will have Hill functions modulate the ongoing **rate** for the flux rather than the flux itself, as seen in the ODEs in ***Equations 3***. Thus Flux = *F*_*h*_*(x*_*i*_*)*x*_*i*_, where *x*_*i*_ is the *i*_*th*_ state, with this functional described by:

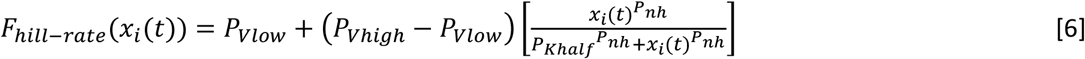

where here we set *P*_*nh*_ = 2 for the shape factor (a higher value such as 4 would provide a sharper transition, a value of 1 gives classic “Michaelis-Menten” kinetics). This enables us to modulate only using the “halfway” transition parameter *P*_*Khalf*_, which should be within the natural range for a given signal. Intuitively, the rate varies from *P*_*Vlow*_ at low values of *x(t)* to *P*_*Vhigh*_ at high values, reaching halfway when *x*_*i*_ *= P*_*Khalf*_. If the user sets *P*_*Vmin*_ = *P*_*Vmin*,,_ we have the special case of a constant rate for that flux. Usually *P*_*Vlow*_ < *P*_*Vhigh*_, and rates increase with signal value, but having the option to have rates be lower with signal value, i.e., *P*_*Vlow*_ > *P*_*Vhigh*_, provides design flexibility (e.g., considered by this author for I3 to R2). For simulations here, the author tended towards caution, using differentials between these extremes that were usually at or under a factor of 2.

### 4.3 Design Considerations for Model Parameters: Principles Underlying Tuning

We’ve seen that the overall model can be viewed as 2 coupled subsystems:

- A closed, 7-state SEI_3_R_2_S **compartmental model** framework, extending the classic SEIRS epidemiological model, intended more for use in public health.
- As the 8^th^ state, a 1^st^-order **pathophysiological (or medical) “placeholder” model** for a collection of hosts that relates to the degree of “not recovered” within hosts and collective clinical outcomes.

For the former, the following 6 principles underly distribution of flow and “storage” of population states for lattice networks:

1. **Conserving “no leak” fluxes (i.e**., **outflux=influx for each arrow):** This implies no loss of population if all fluxes are within a closed system of size N (see **Equations 1**). The exception for states 1-7 is the small “death” outflux from I3.
2. **“Smoothing” unidirectional compartmental flow**: SEIR systems tend to behave like smoothing “low-pass” filters, i.e., “downstream” states tend to exhibit smoother curves (e.g., I1 to I2 to I3).
3. **Summed Outflux/Influx ratio for a compartment: natural dischargers (e.g**., **I’s) and accumulators (e.g**., **R’s)**. For a given compartment, if the sum of influx rates is lower than that for outflux rates, its state tends to eventually drift toward a lower value, i.e., to “push through” population (e.g., the I states). Alternatively, if summed outfluxes are relatively lower, states will tend to accumulate population (e.g., R states, with only gradual outfluxes back toward S).
4. **Values of state variables not governed by gradients**. In bicausal transmission line systems, gradient between states (e.g., voltage, pressure) determine direction (high-to-low) and magnitude of flow, an intuitive concept (e.g., blood vessels). In unidirectional systems without such “backflow” the “downstream” compartments can have higher values than upstream, as will often be the case here (e.g., “accumulating” R states with higher values than I states).
5. **Parallel outfluxes and “path of least resistance” concept**. When there is more than one outflux from a compartment, the proportion of flow distribution depends on the relative values of the ongoing rates, with higher relative rates having proportionately greater outfluxes (e.g., from I3, more to R2 than D if R2 outflux rate is higher).
6. **Rates, with units of /day, intuitively affect temporal dynamics**. While for this higher-order submodel and our formulation one cannot directly think in terms of a “time constant” or “half-life” for any specific flux, it is true that a lower overall rate does imply slower dynamics. It is useful to view rates as a “per day” construct. Note that outfluxes add.

### 4.4 Insights and strategies for model parameters for each of E, I’s, R’s and Nrec

#### E (exposed) Compartment

Exposure is assumed to occur via input-modulated S↔I encounters, with the proportion of E outflux that is directed back to S versus that forwarded onward to I1 depending on the rate ratio. The flux rates associated with the E signal are intriguing to select in the sense that epidemiology and biophysics conflict on timescale. But so long as the E outflux ratio is maintained this will not significantly affect macro-behavior. Specifically, when using traditional SEIR **epidemiological** models, a key reason for expanding from a SIR to a SEIR representation is to produce a **latent time period**: to slow down (i.e., low-pass filter, delay) dynamic flow for a few days via the E state, plus the added rate constant for E can help with data fitting (11).

In contrast, the **biophysics of host-parasite dynamics** involves dynamics on the order of minutes to hours to days, and includes passive viral flow, ACE2 receptor attachment dynamics, intracellular invasion and viral production dynamics, emerging viral load translocation that likely includes advection more than diffusion, and gradual tissue damage through a variety of mechanisms (e.g., due directly to SARS-CoV-2, due to loss of ACE2 function, due to “cross-fire” consequences of immune response), and so on (e.g., 1,2,39-41,61,62).

Both conceptualizations have merit, but each views an “exposure event” differently. It was biophysical analysis dating back to March that led to the recognition that a key transmission mode of viral particles is in the form of **aerosols** (1,2,32-38,63,64), and associated with this, the overriding importance of mask use and of limiting indoor group gatherings (31-38), and less so outdoor gatherings such as protests with high mass use (56).

It took time, perhaps with too much inertia (e.g., WHO (65)), but this is now widely accepted in public health, and now there is ever-expanding epidemiological evidence that mask use remarkably diminishes the risk of exposure events (1,2,34-37). CDC guidance, which continues to evolve in part owing to tension between scientific and political priorities but has recently matured (66), formalizes “exposure” as a window of time in close proximity to an infected person (e.g., “any duration” to 15 min cumulative, without masks). There is much still to learn. From a biophysics perspective, this may be the most important compartment to explore further.

In **epidemiology**, estimates of viral load causing “infection” range from about a hundred to thousands of viral particles, as measured or estimated (67,68). This relates mostly to a PCR testing threshold, which in turn relates to “sensitivity” of the 2 pragmatic testing methods. So, the E to I1 threshold is defined by measurable viral load.

In **biophysics**, even a single inspired airflow could lead to a virion particle successfully binding to ACE2 receptor site and launching the stealth attack via use of the intracellular production machinery. But virions passively “finding” a ACE2 receptor binding site appears to be a somewhat rare biophysical event, given that these particles are part of advective respiratory airflow (and potentially blood flow) and there is only a moderate density of exposed ACE2 transmembrane receptors even on cell types strongly expressing ACE2 (69). SARS-CoV-2 apparently has evolved with improved local electrostatic and hydrophobic binding affinity (70,71), plus enhanced biophysical stability once attached (70), increasing likelihood of a successful attack. Any successful intracellular manufacturing and subsequent export of SARS-CoV-2 virions could be considered as the start of an infection, or it could be clusters of infected cells that have successfully started to translocate, using the respiratory (and circulatory) flow systems. This would differ from the epidemiological perspective.

Actual E→I1 transition dynamics can certainly be less than a few days. Indeed, this author started with E outfluxes on the order of fractions of an hour. The model worked fine, although a byproduct was a strong daily oscillation of the I1 state (even more than in **Figures 4-6**) that was mild but still noticeable by the I2 state.

The version provided here is a compromise **motivated more by epidemiological considerations**, and thus the flux rates are on the order of 2-4/day, i.e., “time constants” from 12-24 hours. Still another criteria was that less than 1% of population should be in the “exposed” state at any given time.

This decision was also justified by another important criteria: the fact that as is normal for SEIR models, the **E→I1 flux launches an irreversible path through the compartmental lattice**, the shortest of which is I1 to R1 (e.g., the time of self-quarantine can encompass E, I1 and part or all of R1 transitions). This implies that we must assume that any I1-stage infection must be enough to **trigger a R1-stage immune response**, even if small.

There are other implicit biophysical assumptions that are part of this approach. For instance, the human **lungs** can be viewed, among other things, as an **aerosol production machine**. Hence why we have defoggers in our cars, why we see the breath of football players in cold weather as it floats away, and why there is so much social media chatter about what to do about fogged glasses while wearing masks. In theory, a maskless person with current immunity (by our threshold criteria) could still be a spreader via the **inhalation-exhalation process**, especially since virions could still briefly take over cell machinery and launch an attack; it just would be gradually turned away by the immune system. We assume that this is negligible, providing no R1→E or R2→E pathways.

To summarize, the E compartment, and really S-E-I1 flux dynamics, can have different interpretations that, with care, still result in similar overall SEIRS performance. But it is ripe for “under the compartmental hood” modeling (see also **Section 3.6**).

#### Infection Dynamics and Stages (I1, I2, I3)

Here we consider a “viral load” sample, measured from one region of tissue, as just one of many possible measures of the stage of an infection. Other indications range from medical images (e.g., lungs, heart) to vital sign-type measurements. The stages used here are designed to relate loosely to meaningful thresholds: significant symptoms (I2-stage) and (ideally) a need for hospitalization (I3-stage).

##### **I1**-stage infection

This can be viewed as the “stealth” stage where “exposure” E has transitioned to infection I1. This mostly asymptomatic stage involves **continued production of new virions** and, importantly, continued **translocation** via the host’s respiratory and/and circulatory systems (likely both). As more ACE2 receptors and the associated cells are compromised, early in this stage an immune response will be launched, starting locally (within cells, immune agents in local tissue) and then systemically (as an agent-based, distributed CAS system). For the majority of hosts the spread will be limited, and a **near-asymptomatic** transition will occur to the R1 stage. Thus, the I1→R1 flux should be larger than the I1→I2 flux, such that roughly 10-20% will proceed to I2. Without extensive SARS-CoV-2 testing, most I1 transitioning to the I1→R1 path will **not be part of “cases”** data.

##### **I2**-stage infection

This is loosely associated with **noticeable symptoms**, which can take manifold forms ranging from fever to difficulty breathing to compromised sense of taste/smell (see CDC advisory for longer list). Some of these reflect viral location, while others such as fever likely relate to systemic effects of the ongoing “battle” between infection spread and immune response. These normally would **classify as “cases”** when adequate testing is available, and for a first approximation the I1→I2 flux can be viewed as **higher than a “daily case” metric**, and much higher than “cases” during Spring 2020 (when even individuals with symptoms were often not tested, and advised to home quarantine). In theory it could be lower than “cases” by Fall 2020 (when through contact tracing and preventative testing “cases” now include many I1-stage positives), but probably not. This is a critical stage, and here we assume that there are **three possible outflux paths**: to R1 (best case scenario), R2 (reflecting a longer process) and the more severe I3 stage. **Medical intervention Input 2** is a dimensionless value over 1.0 and increases the outflux to R1 (and thus tends to mildly lower the I2 trajectory). Code supports increasing this value via slope changes that are tied to improving medical knowledge and drug interventions.

##### **I3**-stage infection

This is designed to be associated with an estimate of health conditions that should (ideally) result in **hospitalization**, assuming health access is available. When viewed this way, since the ratio of hospitalizations/cases has been about 8% (but dropping to ∼4%), it is possible to design the flux from I2 to I3 such that this ratio occurs. The I3→D death flux rate is assumed I3-state dependent. During the I3 stage many **medical interventions** are likely administered, some related to transitioning to R2 but many related to improving recovery (i.e., minimizing Nrec for various tissues and organ systems); the latter is addressed later. **Medical Intervention Input 1**, a dimensionless value over 1.0, both increases the flux to R2 and decreases that to D (death). As medical knowledge has grown, this reflects a decreasing rate of death after hospitalization (less so if overcrowded). As not all make it to hospitals, I3 may be a bit higher than actual hospitalization data.

#### R (resistant to infection) stages

The delineation between the two R states is a simple concept: Available evidence suggests that for mild cases, the “resistant” state will only be on the order of perhaps 3-8 months (48-51). This is consistent with most other members of the coronavirus family (but not SARS-CoV), as well as with a response that involves mostly antibody protein content and a more limited cellular upregulation. The R1→S outflux reflects such dynamics (e.g., 0.01/day). The I1→R1 path is also faster, often resolved within a week. In contrast, R2 is assumed to be associated with a more difficult virus-immune response “battle” and with a larger associated antibody response augmented by greater upregulation of adaptive T-cells and especially B-cells (53-54). Each of I1, I2 and I3 include outfluxes converging to R2. Because of the greater challenges and longer times associated reaching R2 through the I2 and I3 stages, there is more likely residual tissue damage, whether acute or chronic. Thus Nrec is currently assumed to be a function of influxes to R2 and to a lesser extend R2, but presently not to influxes to R1 (though there is evidence for impairment for asymptomatic cases (e.g., 46)).

Notice that we are dramatically simplifying the immune system to a few fluxes. The actual immune response is a natural CAS (27), essentially a distributed information system composed of biophysical agents. SARS-CoV-2 as both a virion and a “community” is highly-tuned to function as an effective parasitic “collective” within the host, taking advantage of its flow systems, a specialized receptor (ACE2), internal production machinery, and an ability to co-exist while awaiting spreading opportunities. Both compartments R1 and R2 could be expanded to have further internal dynamics (e.g., for subpopulations with different risks), and if so, two seems the right number of R’s to start with. Ties to Nrec could then be expanded.

#### General

For all compartments, rate ranges *V*_*low*_ and *V*_*high*_ have the classic units for rate functions, per day, with rates a nonlinear function of the upstream compartment (**Section 4.2**). Usually *V*_*low*_ *< V*_*high*_, but there is flexibility. To linearize a given flux, simply make *V*_*low*_*=V*_*high*_. Since Hill’s *K*_*half*_ has units of normalized subpopulation, values for it can be expected to be lower for E, I2, I3 and R2 than for I1 and R1. In the starter model values have 1 or 1.5 significant digits, and were not extensively tuned, given that the primary contribution of this work is to propose a novel model structure. For cases that involved different risks or special UoD populations, these were mildly adjusted (e.g., Oregon’s lower obesity demographic (case 4), nursing home’s older population (case 6)).

### 4.5 Input Structure and Strategies

Inputs to the model modulate fluxes, nothing more. Inputs fall into two classes: subpopulation encounter modulators, and medical intervention modulators. The latter have already been addressed as part of **Section 4.4**. Here we focus on the former.

#### Input encounter modulation structure and strategy

As noted in **Section 1.2**, this is a “systems engineering” alternative to approaches such as the λ or β modulators. The classic multiplicative “encounter” modulators where included in the state equations (**Equations 3**), but in a concise form that we develop more fully here. The motivation was addressed in **Section 1.2** and **Figure 2**. Here is the form of this mapping:

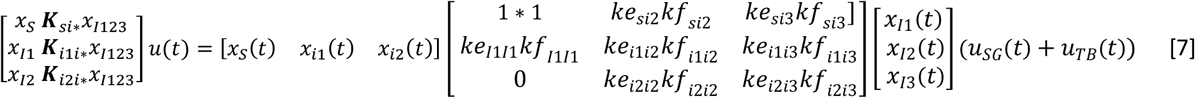

At each time step, the input modulates a product relationship that first includes a sensitivity normalization process. This is “normalized” by viewing the obvious most likely situation of an S-I1 encounter as having a value of 1.0. The summed terms represent the possibility for a S (susceptible) individual to encounter an infective person who may be in any of stages I1, I2 or I3 (top row), for I1 to be worse due to encounters with I1 or I2 or I3 individuals (middle row), and I2 to be worse due to encounters with other I2’s or I3’s. All *ke*’s (effectiveness relative to S↔I1) and *kf*’s (relative frequency relative to S↔I1) that are shown are less than 1.0. Pragmatically, since S is usually over 80% of the population and I1+I2+I3 is rarely over 1%, and I1→I2→I3, the top row dominates. Only the first term of the second row may matter (e.g., I1-I1 interaction during multi-day spreader at social events such as conferences or South Dakota’s Sturgis motorcycle event). The third row is only relevant for special runs involving isolated populations (e.g., nursing home, e.g., as SEIRS-type (72), hospitals).

#### “Instantaneous” Reproduction Ratio

We have de-emphasized R_o_ since it fluctuates (<1 at night, >1 during most of days without strong CMDs). While dominated by S↔I1, there are small S↔I2 and S↔I3 effects (**Equ. 7**), and fluxes are nonlinear. But *u(t)* is “designed” so values around *u=1* are close to 1, and we have this estimate:

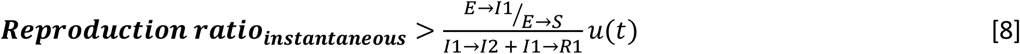

#### Input encounter strategy

Heuristic input design strategies for *u*_*SG*_ and *u*_*TB*_, as introduced in **Figure 2**, involved review of many online resources. Available data for included cases, hospitalizations, and deaths. There were also estimates of location-specific event risks, mask use and social distancing. Some (e.g., CDC, IHME) included downloadable data (so far not used, but then available for any future optimization work). Key resources were:

- CDC, various sites (e.g., https://covid.cdc.gov/covid-data-tracker/#cases_casesper100k),
- New York Times’ “COVID in the U.S.” tracking site, updated daily,
- John’s Hopkins’ Coronavirus Resource Center (https://coronavirus.jhu.edu/),
- Carnegie Mellon’s COVIDcast Real-time COVID-19 Indicators, by Delphi Group (https://covidcast.cmu.edu),
- Georgia Tech’s COVID-19 Event Risk Assessment Planning Tool, (https://covid19risk.biosci.gatech.edu/),
- Univ. of Washington’s IHMU site for COVID, USA (https://covid19.healthdata.org/united-states-of-america?view=mask-use&tab=trend), many model predictions, also mask use and social distancing covariate curves).

### 4.6 Modeling Vaccine Dynamics

Vaccine candidates are rapidly emerging, using a range of strategies that can be loosely classified into 3 types, based on what is delivered via a body tissue (e.g., injection into skeletal muscle, via skin): ***i)*** inactive virions, ***ii)*** viral protein-based, and ***iii)*** mRNA’s intended to use tissue cells (e.g., multi-nucleated skeletal muscle fibers) to create a viral protein. The first 2 have long histories, while the latter is new and is gathering considerable press as it is further along, with several nearing approval. A review of these approaches and logistics issues is beyond our scope, but the model does support a third medical intervention input *u*_*m3*_*(t)* that modulates S→R fluxes.

Based on available data so far (e.g., 48-54) that suggests natural immunity is on the order of a perhaps 3-8 months for mild cases (e.g., R1) to perhaps 4 months to years for more severe (e.g., R2). Multi-year immunity does not appear to be the norm. This is consistent with what could be expected for most members of this class of respiratory coronavirus, and early data for mild COVID-19 cases (48-52), but there is disagreement. Indeed, a thorough recent study tracking 4 markers (antibodies, B cells, 2 T’s) found B memory cells rising for months (53).

The newest approach – mRNA injection – targets mRNA used for spike protein production from mostly multi-nucleated skeletal muscle fibers. Based on this author’s experience with both normal and pathological muscle tissue mRNA transcript portfolios and transcription networks (e.g., 73), as for myo-tissue up/down-regulation is common, this mRNA strategy of “borrowing” the local production machinery for viral spike protein production is clever. But it will be a temporary bolus of production (engineers call this an impulse response), as mRNA half live is short (which helps explain why it is stored cold). Further, the half-life of the subsequent spike protein, which serves no functional purpose, is also probably only a few weeks. Multiple “bolus” injections separated by about 3-4 weeks helps to generate somewhat overlapping and thus a larger (“booster”) response to better trigger a fuller multi-stage immune response (74). While deemed safe, the key is to get well beyond antibody production to considerably upregulate production of an operational community of adaptive B and T-cells, as otherwise “effectiveness” may be high (e.g., over 90%) but only seasonal in most people. Time will tell. Hence the default within the code is for a **user-selected relative weight between S→R1 and S→R2** vaccine-modulated flux paths. If the dream of multi-year immunity emerges, we lower R2→S or add a new R3 with a very low rate.

There are also the **challenges of distribution**, so for present, the current code only supports a slope-change feature for *u*_*m3*_, not pulses, with the slope mostly associated with a second dose for mRNA vaccines. Using slopes also limits the need for an added state for the S→R dynamics. The implicit assumption of seasonal immunity, if true, increases the value of models such as this one, as integrating vaccines and NPI measures such as masks becomes an optimization problem (e.g., the CDC’s Dr. Redford has described measures such as mask use as a form of “vaccine” well into next year; 37). Getting I1<0.1% (see **Figures 4-5**) in the USA may be tough.

### 4.7 Logistics of Running the Model for Various Cases, and Future Directions for Coding

The model is currently provided as well-commented Matlab Live-Script starter file (**ch2_covid19_JWv2.mlx**, contact). By simply changing the case number (integers 0 to 6, 11) on **code Line 27**, the user has access to all the simulations provided in this paper, plus some additional plots and cumulative metrics. To create novel simulations, a user adjusts the P.* **model** parameters and/or P.u.* **input-specifying** parameters, which once run will generate new signals (found under S.*), plus produce plots (which overplots if the Figures from past run(s) are not deleted). It is recommended that changes be done using the code lines under a given switch>case, and that users rename the starter file before making changes, just in case.

There are positives and negatives to providing the model as a Matlab script file. One positive is that the model structure, with all parameters collected under P.*, has been found to be an effective educational tool for disseminating user-centered physiological models, even for users with a limited programming background. No code is hidden within functions, and yet code is conceptually separated (and highly commented) so that it is reasonably clear where parameters can be changed and the purpose of various parts of the code. This can also be a negative, as such ease in changing code also makes it easy to accidently “break” the model.

Other negatives are classic limitations that come with use of **script files versus a modular design** via functions: no structured graphical user interface (GUIs), no numerical integration options (e.g., simple Euler integration is used in starter code, though P.delt = 0.001 days works well here), no advanced tools (systematic sensitivity analysis, numerical optimization). As noted in **Sections 4.4 & 3.2**, the model has strong **cross-sensitivity to certain parameter combinations** associated with outflux rates. This is based on heuristic tuning that was essentially based on the six design rules of **Section 4.3**. This is much stronger than in most physiological models, but common in population ecosystem models where small changes can bifurcate to big effects (e.g., invasive species, climate change). It suggests that classic use of systematic sensitivity analysis, where one parameter is adjusted at a time (essentially taking a partial derivative), may not be as useful an analysis tool anyways.

#### New Versions

Any new versions will use **embedded functions**, which has been the norm for this author for over 35 years and provides more robust (yet refined) control over features and options. This is expected to include both Matlab and Web-based version, both of which build on coding that already exists for other physiological models designed by this author. Coding for the latter integrates HTML/CSS for the user interface with a model converted to Javascript (but will run browser-side, a concern). While an interactive web-based version enables greater public access (although users without adequate browser and memory support may be constrained to shorter-time runs), the key challenge is that this requires the designer to prioritize (and thus limit) the options available to the user. The current hesitancy for releasing such a “cookbook” model framework is that **interface choices** need to evolve to be responsive to **user feedback**, such as: what options to make part of GUIs, what default model parameters to allow a user to change within a GUI before proceeding, what parameters (and ranges) to offer for sensitivity analysis, what forms of targeted input-tuning optimization to offer, how to change the “starter” model for any groups that request newly-tuned parameters for their specific needs, and so on. In any case, the internal structure will include separate functions for parameter selection (with interactive GUI), the model itself, simple sensitivity analysis, quasi-random production options, plotting, and optimization for a few strategic input parameters. Both sensitivity analysis and optimization are computationally intensive, and may crash (e.g., Web) or freeze (e.g., Matlab) without sufficient computing power.

For any future function-structured **Matlab version**, what will be very useful is numerical optimization, both for tuning: ***i)*** model flux parameters so as to improve this or a new version of the models, perhaps based in part on soft “covariate” constraints as used in (11); and ***ii)*** adding the numerical “optimal control” problem of producing input signal trajectories that balance the competing subcriteria of performance (lowering cases, deaths, etc.) versus costs (personal sacrifices, economic, etc.). For the former, there may appear to be a large number of “free” parameters, but in reality there is a reasonably narrow range of viable rate values, as we have reasonably good knowledge of the temporal dynamics of mild and severe cases. Clearly some values should differ if the UoD includes a population with higher obesity, etc., and a subset of parameters can be tuned using available data. The latter optimization approach is quite intriguing and seems to tie to our societal need to balance benefits and costs – a classic engineering optimization formulation. This may include the “feedforward” strategy of addressing how new vaccines gradually entering into the set of options (see also **Figure 10**). Given the author’s experience using the model, it is unlikely that AI numerical methods are necessary, as classic gradient search approaches will likely be successful, as would Matlab’s Global Optimization tools (e.g., particle swarm). Such effort would require access to computational resources beyond this retired emeritus professor’s personal laptop, which was the source for all simulations provided in this paper.

## Data Availability

All data and results provided in the manuscript are available for replication, via code provided by the author.

## Acknowledgements

Access to Matlab was provided by Marquette University, as part of being an emeritus faculty member.

### 6 Appendix: Expanded Simulation Plots for Figure 6 Cases Provided in Default Code

The code is embedded within a descriptive and highly commented live script file, ch2_COVID19_JWv2.mlx, that is available from the author by request (contact via). The runs that follow are screen dumps of Matlab Figure 1, which is one of 3 that automatically plot when a run is complete (Figure 2 gives Hill functionals and rates as a function of key states, similar to “phase plane” and “operating range nonlinearity” plots; Figure 3 gives eight strategic plots in a 2-column format).

All of these are all S.* versus S.t (time vector) plots, where S.* represents a selected signal (e.g., state, input, output, flux). Also provided, selectively, are cumulative metrics from the S.yc container.

**Figure 11:**
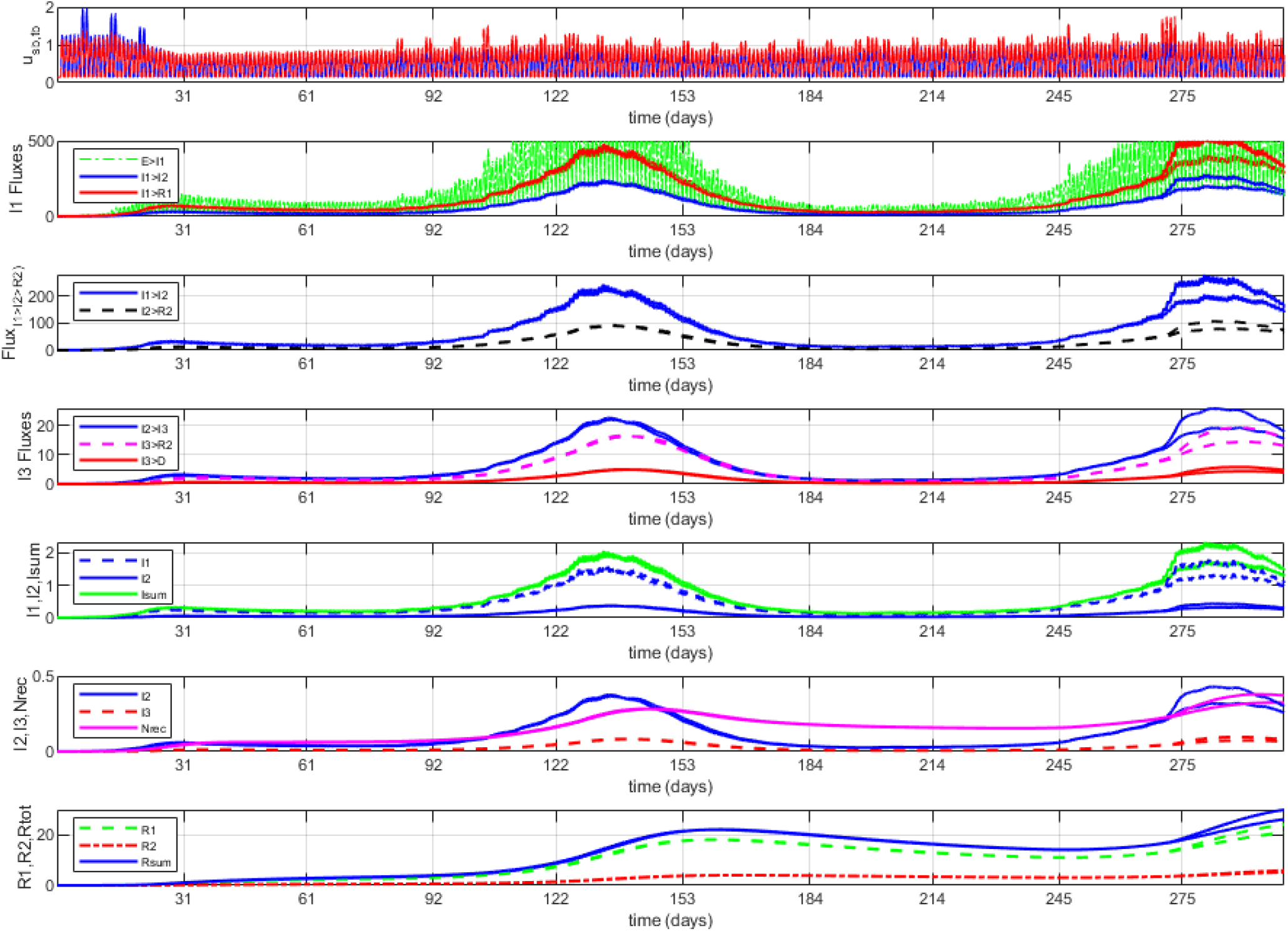
Selected plots (“Figure 1” from Matlab code) from Arizona “starter” simulation (300 days). See **Figure 5** (for USA) for explanation of curves, and for discussion of run setup strategy see **Section 2.5** for Arizona and comments with Matlab script file **Case 2**. Notice the obvious increase during summertime, which was much higher than Wisconsin or Oregon over this time window (**Figure 6**), partly sparked by political events and rapid relaxation of CMDs. There ended up being major hospital capacity challenges, and a high death rate. For roughly Thanksgiving to Christmas, two alternatives are overplotted, with giving a window between differing assumptions on the effects of Thanksgiving (0.6 vs 1.2 pulse) and adherence to CMDs within Arizona society (0.05 difference). Cumulative metrics were stopped at 270 days (i.e., Thanksgiving), and values for these include, given in per 100k (multiply by 73 for Arizona totals): **1)** for fluxes: 44k for E→I1, 14k for I1→I2, 1.4k for I2→I3, 36k for I1→R1, 0.3k for I3→D, and 26k for R1 & R2 →S path; **2)** for signals, at 270 days: 82% for S, 0.85% for I1, 0.2% for I2, 0.04% for I3, 13% for R1, and 3.5% for R2.

**Figure 12:**
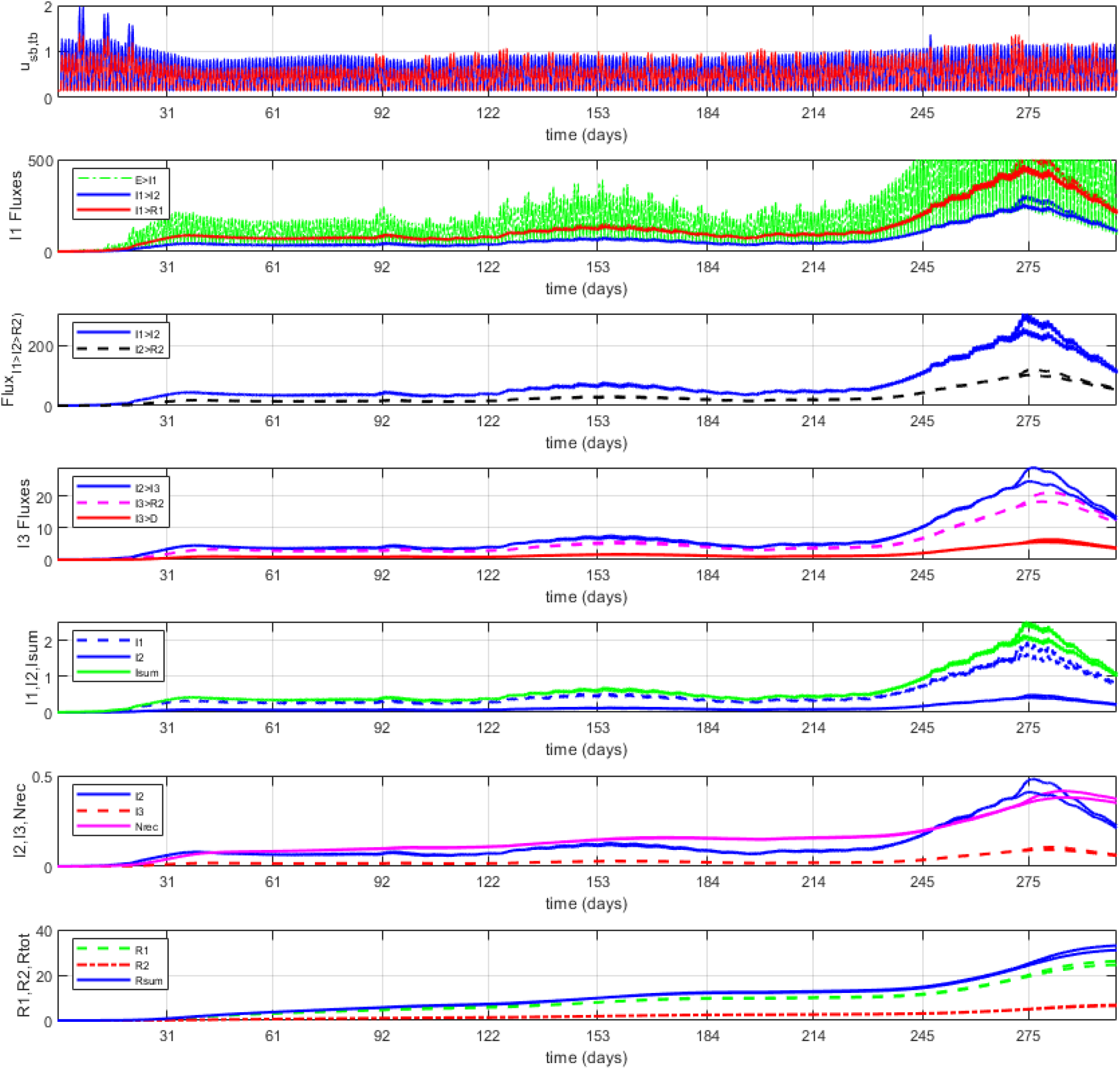
Selected plots (“Figure 1” from Matlab code) from Wisconsin “starter” simulation (300 days). See **Figure 5** (for USA) for explanation of curves, and for discussion of run setup strategy see **Section 2.5** for Wisconsin and comments with Matlab script file **Case 3**. The model was slightly adjusted for the higher obesity rate (*V*_*high*_ from I’s to R1’s 5-10% lower (slower average recovery), I3→D ∼10% higher). Notice the fluctuations throughout Spring-Summer, and then the obvious increase during the Fall that is well above Arizona and Oregon (**Figure 5**). For December (through roughly Christmas), two alternatives are overplotted, both predicting a drop (as S lowers below 80% and R’s cross 20%), with differing assumptions on the effects of Thanksgiving)0.6 vs 1.2 pulse) and adherence to CMDs within Wisconsin society (0.05 slope change). Cumulative metrics are given at 270 days (i.e., to Thanksgiving), and values for these include, given per 100k (multiply by 58.3 for Wisconsin totals): **1)** for fluxes: 45k for E→I1, 15k for I1→I2, 1.4k for I2→I3, 36k for I1→R1, 0.032k for I3→D, and 20.7k for R1 & R2 →S path; **2)** for signals, at 270 days: 76% for S, 1.4% for I1, 0.37% for I2, 0.08% for I3, 17.7% for R1, and 4.5% for R2.

**Figure 13:**
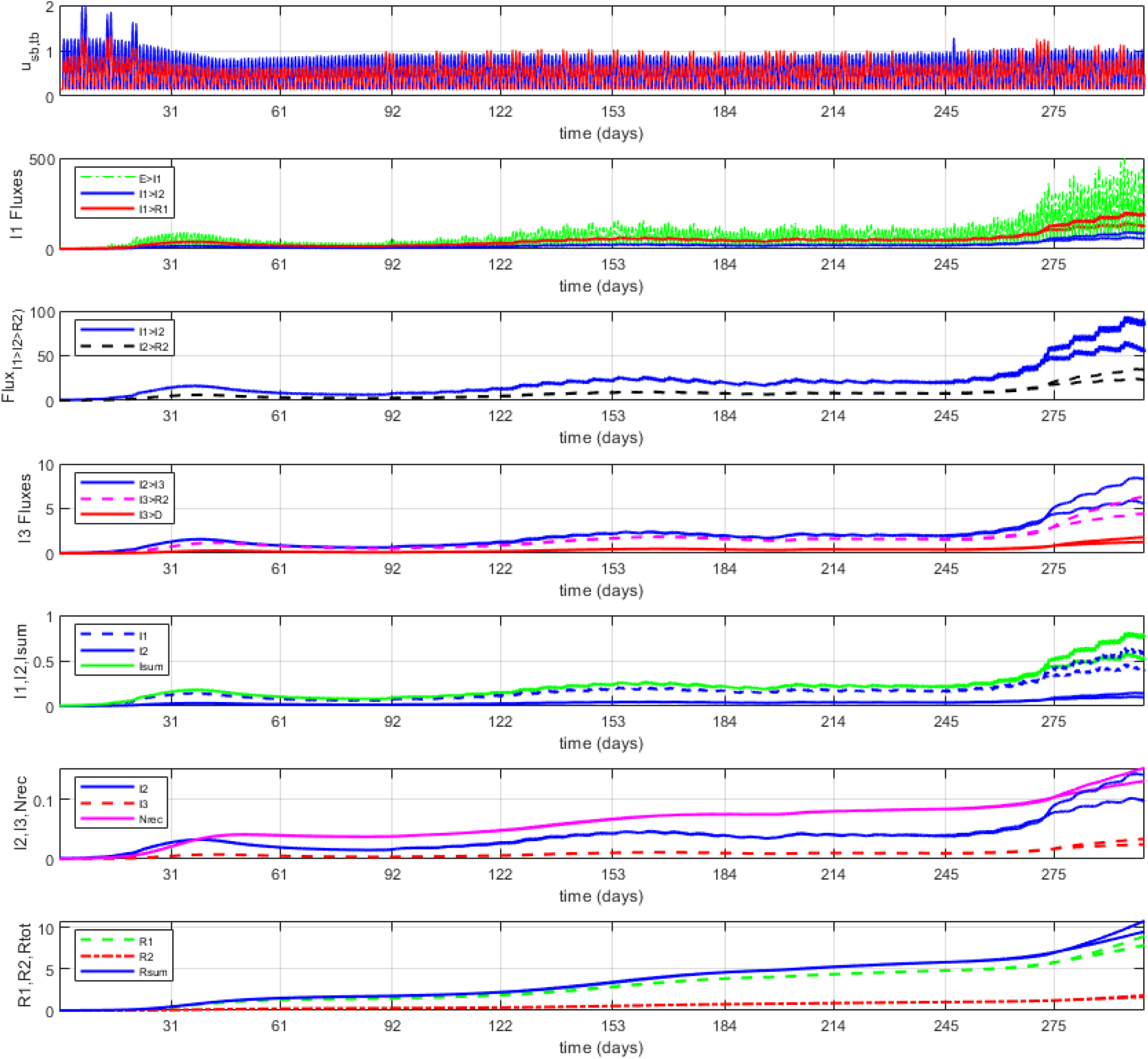
Selected plots (“Figure 1” from Matlab code) from Oregon “starter” simulation (300 days). See **Figure 5** (for USA) for explanation of curves, and for discussion of run setup strategy see **Section 2.5** for Oregon and comments within Matlab script file **Case 4**. The model was slightly adjusted for the relatively lower state obesity rate (*V*_*high*_ and *V*_*low*_ from I’s to R1’s 5-10% higher (faster average recovery), I3→D ∼5% lower). While there have been a few increases that looked like they could lead to an exponential rise, through October this did not happen, and CMDs have remained largely in place, especially for the metro Portland region. Increases in cases, hospitalizations and deaths have risen during November have led to recent CMDs, and time will tell, with two alternatives overplotted with differing assumptions on the effects of Thanksgiving and adherence to CMDs within Oregon society. Cumulative metrics are given at 270 days (i.e., to Thanksgiving), and values for these include, given per 100k (multiply by 43 for Oregon totals): **1)** for fluxes: 14.6k for E→I1, 4.2k for I1→I2, 0.41k for I2→I3, 12.3k for I1→R1, 1.9k for I2→R1, 0.008k for I3→D, and 7.6k for R1 & R2 →S path; **2)** for signals, at 270 days: 93% for S, 0.26% for I1, 0.06% for I2, 0.014% for I3, 5.4% for R1, and 1.1% for R2. Notice that the model predicts that Oregon didn’t even cross 5% “resistant” (*R*_*tot*_) until September, and approaches December with only about 7% (and growing), likely to cross 10% before the end of the year but still under half of those predicted for Arizona and Wisconsin, but on a relatively steeper upward trajectory.

